# CXCR4 inhibition in human pancreatic and colorectal cancers induces an integrated immune response

**DOI:** 10.1101/2020.07.08.20129361

**Authors:** Daniele Biasci, Martin Smoragiewicz, Claire M. Connell, Zhikai Wang, Ya Gao, James Thaventhiran, Bristi Basu, Lukasz Magiera, Isaac T. Johnson, Lisa Bax, Aarthi Gopinathan, Christopher Isherwood, Ferdia Gallagher, Maria Pawula, Irena Hudecova, Davina Gale, Nitzan Rosenfeld, Petros Barmpounakis, Elizabeta Cristina Popa, Rebecca Brais, Edmund Godfrey, Fraz Mir, Frances Richards, Douglas T. Fearon, Tobias Janowitz, Duncan Jodrell

## Abstract

Inhibition of the chemokine receptor CXCR4 in combination with blockade of the PD-1/PD-L1 T cell checkpoint induces T cell infiltration and anti-cancer responses in murine and human pancreatic cancer. Here we elucidate the mechanism by which CXCR4 inhibition effects the tumor immune microenvironment. In human immune cell-based chemotaxis assays, we find that CXCL12-stimulated CXCR4 inhibits the directed migration mediated by CXCR1, CXCR3, CXCR5, CXCR6, and CCR2, respectively, chemokine receptors expressed by all the immune cell types that participate in an integrated immune responses. Inhibiting CXCR4 in an experimental cancer medicine study by one-week continuous infusion of the small molecule inhibitor, AMD3100 (plerixafor), induces an integrated immune response that is detected by transcriptional analysis of paired biopsies of metastases from patients with microsatellite stable colorectal and pancreatic cancer. This integrated immune response occurs in three other examples of immune-mediated damage to non-infected tissues: rejecting renal allografts, melanomas clinically responding to anti-PD1 antibody therapy, and microsatellite instable colorectal cancers. Thus, signaling by CXCR4 causes immune suppression in human PDA and CRC by impairing the function of the chemokine receptors that mediate the intratumoral accumulation of immune cells.

**Statement of significance:** Continuous infusion of AMD3100, an antagonist of the chemokine receptor, CXCR4, induces an integrated anti-cancer immune response in metastases of patients with microsatellite stable pancreatic and colorectal cancer that is predictive of response to T cell checkpoint inhibition.

## Introduction

T cell checkpoint antagonists that target the regulatory membrane proteins on T cells, CTLA-4 and PD-1, have demonstrated the therapeutic potential of the immune system in cancer. Clinical responses, however, have been limited to subsets of patients with certain cancers (1–4). Lack of cancer cell antigenicity (5), dysfunction of cytotoxic CD8+ T cells (6), and systemic immune modulation (7,8) have been some of the potential explanations for resistance of these cancers to T cell checkpoint inhibitors. A more general immunological principle in which mesenchymal cells may control the immune response to immunogenic epithelial tissues should also be considered (9).

The presence of tertiary lymphoid structures (TLSs) in human adenocarcinomas correlates with better long-term clinical outcome and clinical response to T cell-checkpoint inhibitors (10–13), suggesting that organized intratumoral lymphoid structures promote anti-tumor immune reactions. Mesenchymal stromal cells organize B and T cells in both secondary and tertiary lymphoid structures mainly by producing chemokines: CCL19 and CCL21 from fibroblastic reticular cells (FRCs) recruit CCR7-expressing lymphocytes and dendritic cells (DCs), and CXCL13 from follicular dendritic cells (FDCs) attracts CXCR5-expressing T and B cells (14). Notably, these two stromal cell types develop from an embryonic precursor that expresses the membrane protein, Fibroblast Activation Protein-*α* (FAP), and may be developmentally related to the FAP-expressing fibroblastic stromal cell type that resides in solid tumors, which is termed the cancer-associated fibroblast (CAF) (15–18). CAFs also affect the trafficking of lymphocytes by producing a chemokine, CXCL12 (19), but in a manner that opposes the effects of lymphoid tissue stromal cells, by suppressing the intra-tumoral accumulation of T cells (20). Indeed, continuous inhibition of CXCR4 in a mouse model of pancreatic cancer leads to T cell infiltration of the tumors and results in response to anti-PD-L1 antibody administration (20). Therefore, whether the tumor stroma supports or suppresses immune activation may depend on the relative contributions of these related stromal cell types. A pre-dominance of CAFs would suppress local immunity, whereas the presence of FRCs and FDCs and the development of TLSs would enhance intratumoral immunity. This concept has been supported by pre-clinical studies in which immune control of tumor growth in mice occurred after FAP+ CAFs were conditionally depleted (18,20).

Here we report the results from a proof of concept experimental medicine study in which we tested the immunological consequences of inhibiting CXCR4 in patients with cancers that have historically resisted immunotherapy, micro-satellite stable (MSS) colorectal cancer (CRC) or pancreatic ductal adenocarcinoma (PDA). We report that continuous administration for one week of AMD3100 (plerixafor, Mozobil^™^), a small molecule inhibitor of CXCR4, promotes an integrated immune response in metastatic lesions from these patients.

## Results

### Colorectal and pancreatic cancer cells display a CXCL12 “coat”

In murine PDA tumors, cancer cells display a “coat” of CXCL12, the chemokine that FAP+ CAFs produce to mediate their immune suppressive activity (20). We assessed whether such a CXCL12-coat is displayed on human PDA or CRC cancer cells by examining tumor tissue microarrays. Fluorescently labeled anti-CXCL12 antibodies stained the KRT19-expressing cancer cells in formalin fixed paraffin embedded (FFPE) tissue sections of human PDA and CRC (Fig. 1).

**Figure 1.**
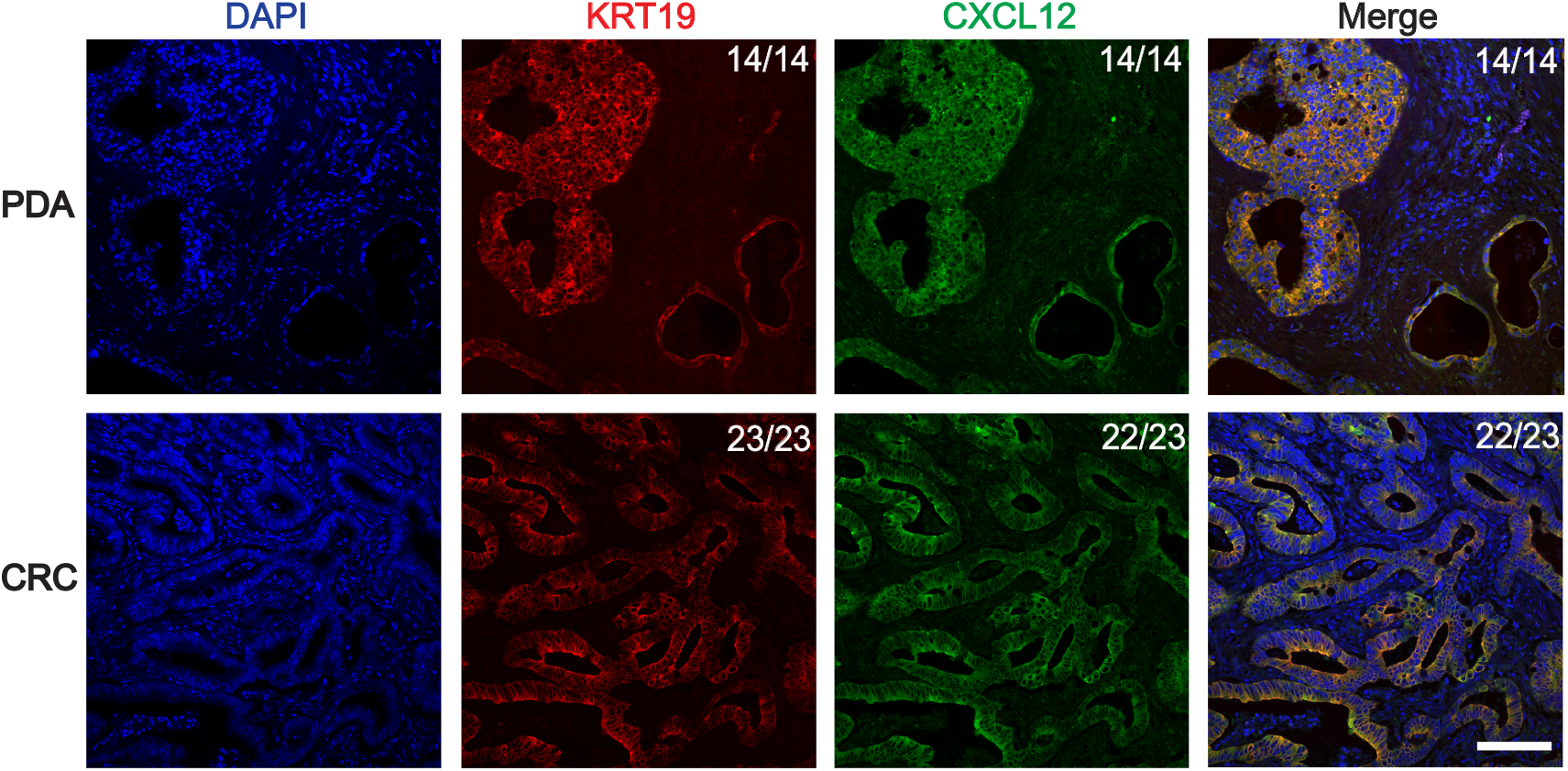
The CXCL12-coat of human pancreatic and colorectal cancer cells. Sections of human pancreatic (PDA) and colorectal (CRC) adenocarcinoma were stained with fluorescent antibodies to CXCL12, and to KRT19 to reveal cancer cells. The ratios shown in the top right corners of the photomicrographs indicate the frequency of the observed staining relative to the total number of independent tumors that were assessed. Scale bar, 50 µm.

### CXCL12 stimulated CXCR4 inhibits the directed migration of human immune cells mediated by chemokine receptors

We assessed whether CXCL12 stimulation of CXCR4 altered the trafficking of human immune cells by performing *in vitro* transwell migration assays using Boyden chambers. We generated human cell lines that co-expressed CXCR4 with the chemokine receptors that mediate the directed migration of innate and adaptive immune cells: CXCR1 in Jurkat-CXCR4/CXCR1 cells, CXCR3 in HSB2DP-CXCR4/CXCR3 cells, CXCR5 in Raji-CXCR4/CXCR5 cells, CXCR6 in Jurkat-CXCR4/CXCR6 cells, and CCR2 in Molm13-CXCR4/CCR2 cells, respectively (Fig. 2 and Fig. S1). These chemokine receptors and their respective chemokines mediate the directed migration of neutrophils (CXCR1; CXCL8), T cells, dendritic cells, and NK cells (CXCR3; CXCL9 and CXCL10), B cells (CXCR5; CXCL13), tissue-resident memory T cells (CXCR6; CXCL16), and monocytes and macrophages (CCR2; CCL2) (Fig. 2). Including CXCL12 in the upper chamber of the Boyden two chamber assay inhibited the migration of the human immune cells co-expressing CXCR4 with each one of the five other chemokine receptors towards its relevant chemokine in the lower chamber. The inhibition was dependent on CXCR4 expression (Fig. S1) and was unidirectional, in that stimulating the relevant immune cell lines with their respective chemokines for CXCR1, CXCR3, CXCR5, CXCR6 or CCR2 did not abolish the CXCR4-mediated chemotactic response to CXCL12 (Fig. 2). Incubating HSB2DP-CXCR4/CXCR3 cells with CXCL12 followed by removal of the chemokine restored the ability of cells to migrate in response to a CXCL10 gradient (Fig. S2).

**Figure 2.**
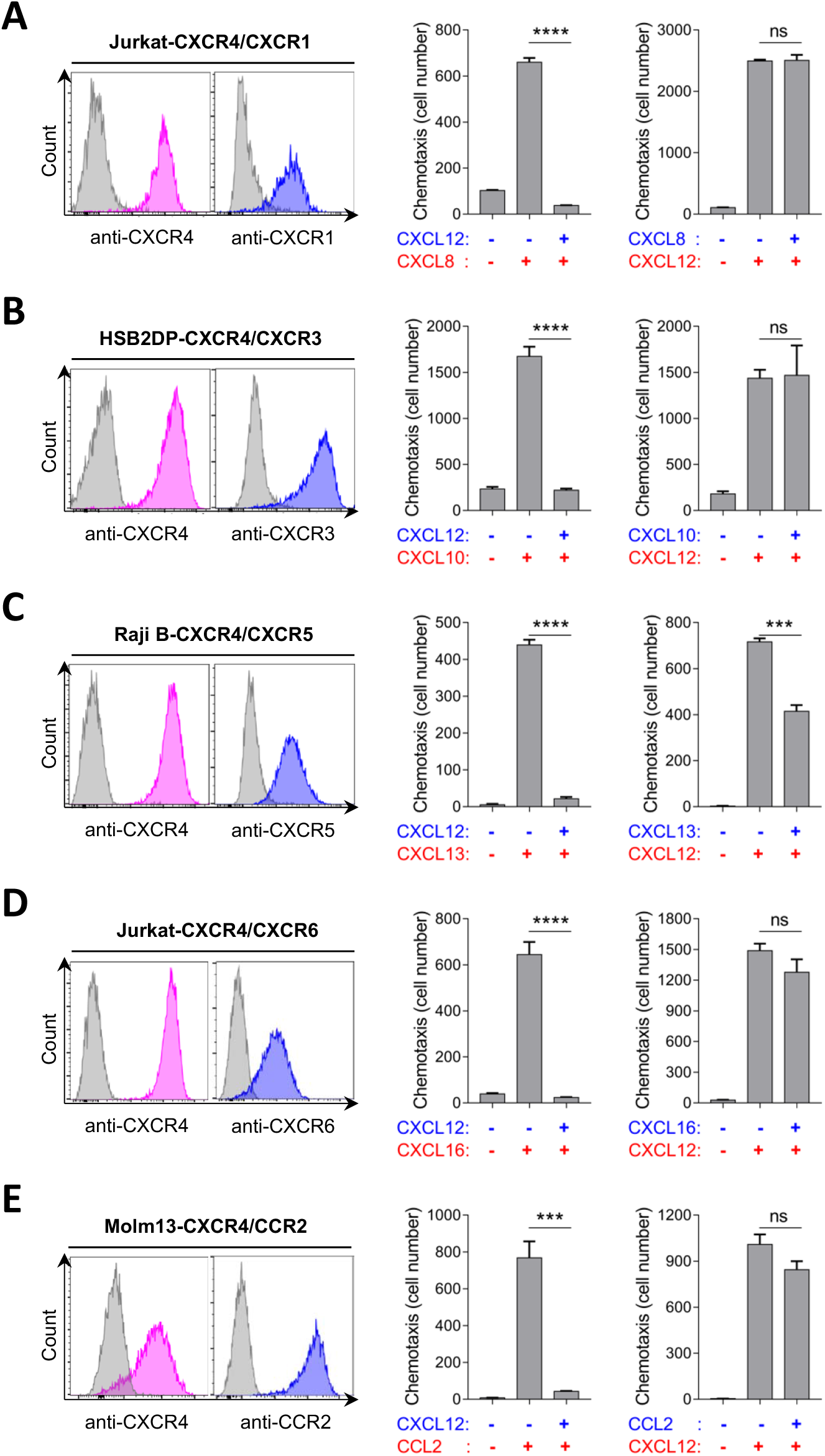
The effect of CXCL12-stimulated CXCR4 on chemokine receptor-mediated migration of human immune cells. Left panels: The co-expression of CXCR4 with (**A**) CXCR1, (**B**) CXCR3, (**C**) CXCR5, (**D**) CXCR6, and (**E**) CCR2 on human immune cell lines was evaluated by flow cytometry after staining with antibodies specific for the relevant chemokine receptors. Gray peaks indicate isotype controls. Middle panels: The effect of CXCL12-stimulation of CXCR4 on the chemotactic responses of (**A**) CXCR1-co-expressing Jurkat T lymphoblastoid cells to CXCL8, (**B**) CXCR3-co-expressing HSB2DP T lymphoblastoid cells to CXCL10, (**C**) CXCR5-co-expressing Raji B lymphoblastoid cells to CXCL13, (**D**) CXCR6-co-expressing Jurkat T lymphoblastoid cells to CXCL16, and (**E**) CCR2-co-expressing Molm13 monocytoid cells to CCL2 was assessed by including CXCL12 in the upper chamber (blue) and the other chemokines in the lower chamber (red) in the Boyden two-chamber assay. Right panels: The chemotaxis assays were performed with the five cell lines when the placement of the chemokines in the Boyden chambers was reversed. Bar diagrams display mean and standard error of the mean. Statistical analysis by student’s t test: ***, p<0.001; ****, p<0.0001; ns, not significant.

### AMD3100 suppresses CXCL12-stimulated inhibition of other chemokine receptors

In previous murine studies a continuous plasma concentration of 2 *μ*g/ml (4 *μ*M) AMD3100 unmasked anti-PDA immunity and led to reduced tumor growth rates and synergy with anti-PD-L1 treatment (20). We thus examined the effect of AMD3100 in chemotaxis studies across this range of drug concentration in these human cell lines. AMD3100 fully inhibited the CXCR4-mediated chemotactic responses of all immune cell lines (Fig. 3). The CXCR4 inhibitor also fully reversed the inhibition by CXCL12 of the chemotactic functions of CXCR1 and CXCR6 on the CXCR4-expressing Jurkat cells (Fig. 3). The functions of CXCR3, CXCR5 and CCR2 were only partially restored, which correlated with the inhibitory effects of AMD3100 on CXCR3-, CXCR5-, and CCR2-mediated chemotaxis in the absence of CXCL12 (Fig. S3). This inhibitory effect of AMD3100 may be caused by partial agonism of CXCR4, which has been reported (21). Since CXCR1, CXCR3, CXCR5, CXCR6 and CCR2 mediate the trafficking of neutrophils, T cells, NK cells, DCs, B cells, tissue-resident memory T cells and monocytes, these observations suggest that AMD3100 may alter the trafficking of multiple immune cell types within tumors, thereby inducing an integrated immune response to the cancer cells. We tested this hypothesis in an experimental medicine study.

**Figure 3.**
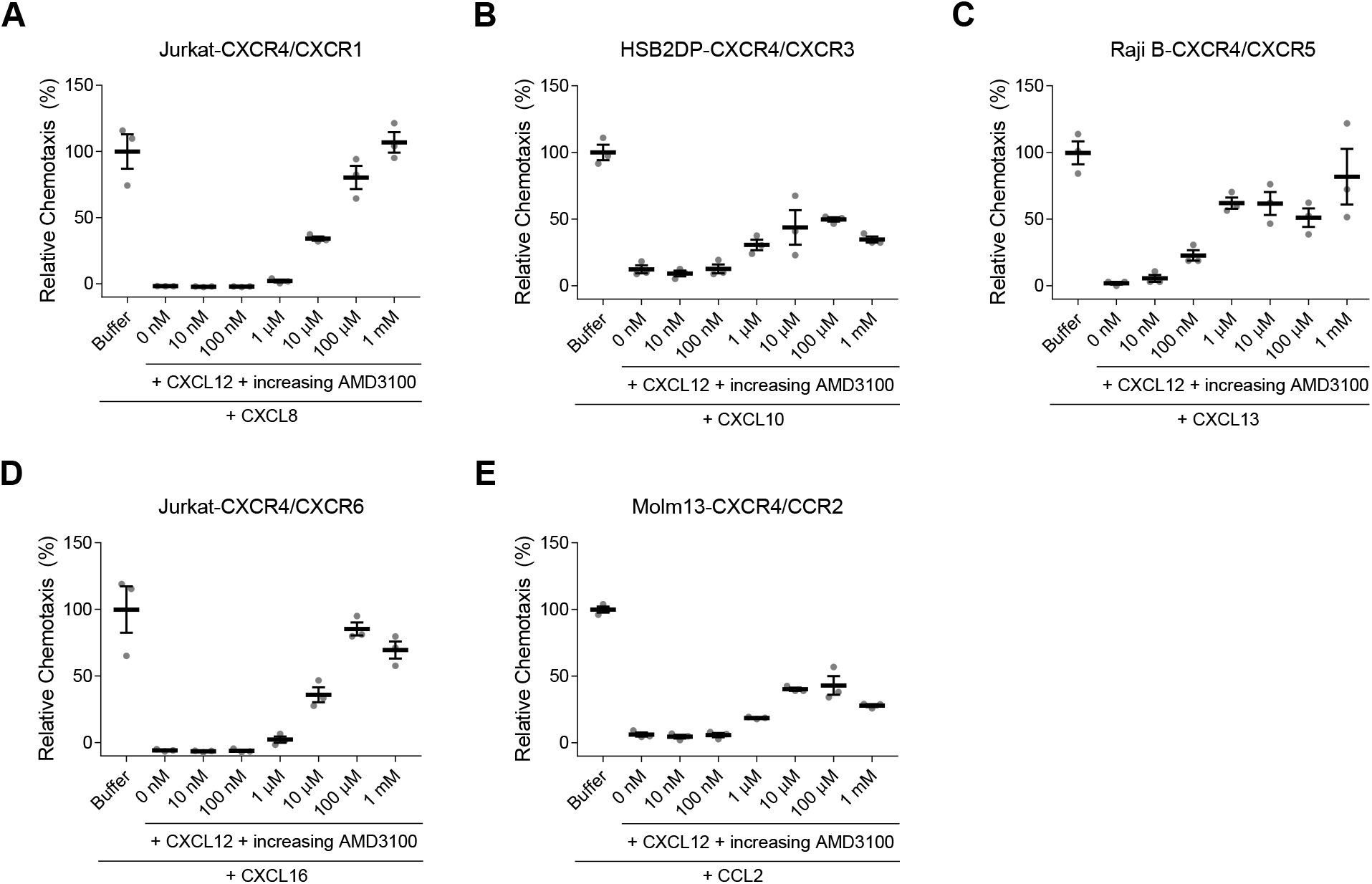
Inhibition by AMD3100 of the suppression mediated by CXCL12-stimulated CXCR4 of the function of other chemokine receptors. **Panels A-E:** The chemotactic responses were assessed of the dual chemokine receptor-expressing human immune cells to the relevant chemokines in the absence or presence of CXCL12, with increasing concentrations of AMD3100. The results are presented as percent of the chemotactic response in the absence of CXCL12 and AMD3100. The mean and standard error of the mean are indicated.

### Experimental medicine study of continuous AMD3100 infusion: Study design, recruitment and patient characteristics

We targeted the CXCL12/CXCR4 interaction using AMD3100 in an experimental medicine study of the immunological role of CXCR4 signaling in patients with MSS CRC and MSS PDA (NCT02179970). AMD3100 has a plasma half-life of approximately 8 hours. To achieve continuous inhibition of CXCR4, as has been recommended for other chemokine receptors (22), AMD3100 was delivered by continuous intravenous infusion for 7 days with the target steady-state plasma concentration being approximately 2 *μ*g/ml (4*μ*M). We assessed the pharmacokinetics, pharmacodynamics, and intratumoral immunological changes during treatment using serial blood tests, clinical imaging modalities, and investigations from paired biopsies taken prior to and at the end of the AMD3100 infusion (Fig. S4 and Fig. S5A).

We enrolled 26 patients at two centers, 24 at the Cambridge University Hospitals NHS Foundation Trust and two at Weill Cornell Medicine/New York Presbyterian Hospital. The patient eligibility criteria are shown in the Materials and Methods section. The characteristics of all enrolled patients are summarized in Table 1. On histopathological review, one patient was found to have predominantly neuroendocrine cancer cells in the biopsy tissue and this patient was therefore excluded from all analyses other than the safety and pharmacokinetic analysis. The remaining 25 patients had treatment-refractory, histologically confirmed MSS PDA (n=10) or MSS CRC (n=15). An important inclusion criterion was the presence of a baseline lymphocyte count above the lower limit of normal (1.0 × 10^9^/L) at screening, because of concerns relating to adequate immune status and resolution of immunosuppression after previous chemotherapy.

**Table 1.** Patient Characteristics.

| **Table 1. Patient Characteristics** |  |
| --- | --- |
| **Characteristic** | **N=26** |
| Age - yr |  |
| Mean | 66 |
| Range | 50-76 |
| Female sex – no (%) | 9 (35) |
| Histology - no. (%) |  |
| Pancreatic adenocarcinoma | 10 (38) |
| Colorectal adenocarcinoma | 15 (58) |
| Neuroendocrine cancer<sup>¶</sup> | 1 (4) |
| ECOG - no. (%) |  |
| 0 | 9 (35) |
| 1 | 17 (65) |
| Lymphocyte count (10<sup>9</sup>/L) |  |
| Median | 1.41 |
| Range* | 0.82-2.31 |
| Albumin < 35 g/L - no. (%) | 13 (50) |
| Sites of metastasis >2 - no. (%) | 16 (62) |
| Prior lines of chemotherapy- no. (%) |  |
| 0 | 1(4) |
| 1 | 1(4) |
| 2 | 16 (62) |
| ≥3 | 8 (31) |
| <sup>¶</sup> On central review of research biopsies, pathology consistent with neuroendocrine pancreatic cancer, excluded from later analysis<br>*1 patient had a low count at enrolment that normalized on day 1 pre-infusion |  |

Twenty-four patients with CRC or PDA were treated with AMD3100 (one registered patient did not commence study drug, because of a disease related adverse event (AE)): 17 in the dose escalation phase (two, PDA; 15, CRC) and seven further patients with PDA in the dose expansion phase. We confirmed the presence of the CXCL12-coat in all patients enrolled in the dose escalation phase who had evaluable tissue (Fig. S6).

### Pharmacokinetic and toxicity results

The first dose level of AMD3100 was an iv infusion at a rate of 20 µg/kg/hr, with subsequent patients enrolled at dose cohorts of 40, 80, and 120 µg/kg/hr, using a 3+3 design. There were no Dose Limiting Toxicities (DLTs) identified in the 20, 40 and 80 µg/kg/hr dose cohorts, but 2 patients experienced DLTs at the 120µg/kg/hr dose (Table S1). One patient had a vasovagal reaction (grade 3) in the context of pain shortly after the Day 8 biopsy and prior to completion of the AMD3100 infusion. One patient who had peritoneal disease developed severe abdominal pain (grade 3), hypotension (grade 3), and a vasovagal reaction (grade 3) on day 2 of the infusion. Symptoms resolved within 24 hr after discontinuing the drug, medications for pain control, and IV fluids. Continuous infusion of AMD3100 has been reported to be associated with vasovagal reactions (23), and these events were classified as DLTs. A complete list of graded AEs is included in the supplemental material (Table S2).

In the trial of continuous IV infusion of AMD3100 for one week in 40 patients with HIV, a single patient experienced premature ventricular contractions (23). Thus, in the current study, all patients were admitted for the initial 72 hrs of the AMD3100 infusion for continuous cardiac telemetry monitoring, and Holter monitoring thereafter. No cardiac rhythm disturbances were identified at the 80 □g/kg/hr infusion rate chosen for the expansion phase. Minor changes (Table S2) were only possibly drug related, and there were no cardiac AEs that required drug interruption and all resolved without sequelae. Therefore, hospital-based telemetry is not indicated in future studies using this infusion protocol and static ECGs and ambulatory Holter monitoring should provide sufficient cardiac monitoring.

The dose rate of 80 µg/kg/hr yielded the target plasma level of ∼2 µg/ml (4µM), and was chosen for the expansion cohort which resulted in a mean steady state plasma concentration of 2.3 µg/ml (SD ± 0.9 µg/ml) AMD3100 in patients enrolled at this dose rate (Fig. S5B, Table S3). This infusion rate was overall well tolerated (Table S2).

### Pharmacodynamic and clinical results

In accordance with the well characterized biological role of CXCL12/CXCR4 ligation for the retention of hematopoietic stem cells and immature leukocytes in the bone marrow (24), CD34^+^ and other leukocyte populations were elevated during the period of AMD3100 infusion. These changes had almost completely resolved by day 28 of the study, 20 days after discontinuation of the infusion (Fig. S7). By conventional CT scanning on day 20-24, no complete or partial responses were identified in 23 evaluable patients. Thirteen patients (57%) had stable disease and 10 (43%) disease progression. Paired PET-CT scans were evaluable in 19 participants (12 escalation phase, 7 expansion phase). Of these, 11 participants had CRC and 8 PDA. Clinically significant (defined as delta Standardized Uptake Values mean weighted average (SUV MWA) ≥ 30%) changes were seen in 2 participants. Both were patients with CRC (treated at 40 µg/kg/hr) and had a ≥30% increase in SUV MWA (71% and 32%).

### Immunological analysis of metastatic tissue from pre-treatment and on-treatment biopsies

We performed immunological analyses on all patients who had sufficient sample material for comparative histopathological and RNA studies (PDA n=4; CRC n=10) (Fig. S4). We first sought histological evidence of an immunological response to CXCR4 inhibition by determining the frequency of CD8^+^ T cells in paired pre- and end-of-treatment biopsies of metastatic lesions. FFPE tissue sections were stained with antibodies specific for CD8*α* and keratin (pan-CK), respectively (Fig. 4A). There was a significant increase in the number CD8^+^ T cells in the pan-CK^+^ cancer cell areas after treatment with AMD3100 (Fig. S8A). Frozen samples of different biopsy passes of the same metastatic lesions were subjected to bulk RNA-Seq analysis. The CD8a mRNA levels were significantly upregulated after AMD3100 administration (Fig. S8B), and significantly correlated with the histopathologically determined CD8^+^ T cell frequencies (Fig. 4B). This validation of the quantitative RNA-Seq transcriptomic set justified the further analysis of this comprehensive source of immunological data.

**Figure 4.**
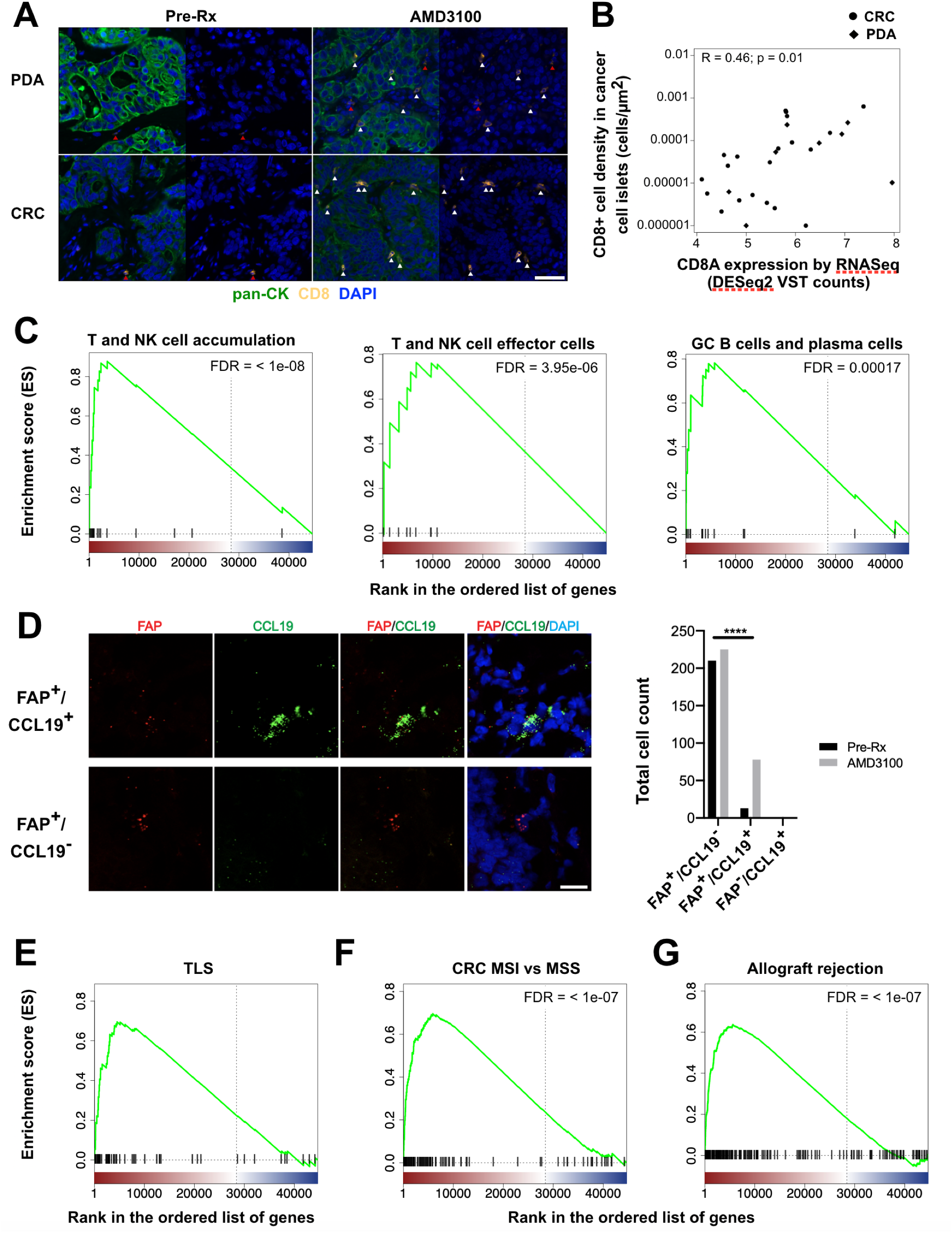
The immunological effects in human CRC and PDA of treatment with AMD3100. Paired biopsy tissues were obtained from the same metastasis in each patient before (pre-Rx) and after seven days of continuous infusion of AMD3100. (**A**) Tissue sections were stained with fluorescent antibodies to pan-keratin (pan-CK) to reveal cancer cells, and to CD8 to reveal cytotoxic T cells. White arrowheads designate CD8^+^ T cells within cancer cell islets, and red arrowheads designate CD8^+^ T cells outside of cancer cell islets. (**B**) The presence of CD8^+^ T cells within cancer cell islets, assessed by staining with anti-CD8 antibody, correlates with the CD8A mRNA levels, assessed by RNAseq analysis, in tissues obtained from different pass biopsies of the same metastatic lesions. (**C)** Immunological gene sets that identify T and NK cell accumulation, T and NK cell effector cells, activated B cells (germinal center B cells) and plasma cells are enriched in genes upregulated after treatment with AMD3100. (**D**) The expression of CCL19 and FAP in sections from paired biopsies was analyzed by fluorescent in situ hybridization using specific probes for mRNA of FAP and CCL19. The total counts of FAP^+^/CCL19^+^ cells, FAP^+^/CCL19^-^ cells, and FAP^-^/CCL19^+^ cells are displayed. (**E**) The enrichment analysis for a TLS gene set (13) is shown. (**F** and **G**) Enrichment analyses are shown for those genes that (**F**) are differentially expressed in rejecting compared to non-rejecting kidney allografts (28, 29) and (**G**) MSI compared to MSS CRC (30). (**A-G**) n=14 comprising of PDA (n=4) and CRC (n=10). Scale bar, 50 µm. Statistical comparisons by Spearman’s rank correlation test (**B**), and by Fisher’s exact test (**D**): ****, p<0.0001.

By enrichment analysis of the RNA-Seq data, we found that continuous inhibition of CXCR4 by infusion of AMD3100 induced intra-tumoral T and NK cell accumulation and activation (Fig. 4C) and also induced an activated B cell response. Eighteen of the 100 most differentially expressed genes were derived from the B cell lineage. The upregulated expression of transcripts encoding the J chain, heavy chain constant regions, and TNFRSF17, when taken together with the more modest increased expression of MS4A1 (CD20), CD19, TNFRSF13B and TNFRSF13C suggests that this response is more a consequence of plasma cell differentiation than an accumulation of B cells (Table S4). A plasma cell transcriptional signature in breast and lung adenocarcinomas has been shown to correlate with improved survival (25). This AMD3100-induced T and B cell response was accompanied by transcriptional evidence for the development of TLSs (Fig. 4D and E, Table S4). The restriction of CCL19 mRNA, which is expressed only by FRCs (26), to tumor stromal cells that were FAP^+^, a marker not only of CAFs but also of FRCs (15,27), was demonstrated by fluorescent *in situ* hybridization (FISH). The FAP^+^ cells expressing CCL19 increased significantly from 5.8% to 25.7% (Fig. 4D), suggesting either that CAFs were differentiating to FRCs within the tumor microenvironment, or that FRCs were being recruited from a source outside the tumors.

### The INTegrated Immune REsponse (INTIRE) and immune-mediated damage to non-infected tissues

The similarity of the AMD3100-induced transcriptional changes to those that characterize tumors with TLSs (Fig. 4E), micro-satellite instable (MSI) CRC (Fig. 4F), and rejecting renal allografts (Fig. 4G) suggested that the inhibition of a single chemokine receptor, CXCR4, may induce an integrated immune reaction that is characteristic of non-infected, immunogenic tissues. To assess the integrated immune response, we developed the INTIRE gene signature which was defined by 194 genes that together identify nine components of innate and adaptive immunity (Table S5): Monocyte/Macrophage/DC/Antigen Presentation, T and NK Cell Accumulation, T and NK Effector Cells, Chemokines and Chemokine Receptors, Activated B Cells (Germinal Center B Cells) and Plasma Cells, Stromal/FRC/TLS, Type I/III Interferon Response, and Endothelial Cells (blood and lymphatic). AMD3100 treatment of MSS PDA and MSS CRC patients induced the INTIRE gene signature (Fig. 5A). Upregulation of the INTIRE gene panel also was associated with decreased expression of E2F target genes and other genes involved in the G2M checkpoint, possibly indicating a reduction in replicating cancer cells in the tumor. The INTIRE gene signature also distinguished between rejecting and non-rejecting renal allografts in two studies (28,29), a prototypical example of immunologically mediated damage to non-infected tissue. Relative to MSS CRC, MSI CRC demonstrated the INTIRE gene signature (Fig. 5A) (30,31), but, in contrast to the effects of inhibiting CXCR4 in MSS CRC and MSS PDA, MSI CRC did not exhibit decreased Cell Cycle gene expression (Fig. 5A). The INTIRE gene signature was also present in tumors from longer surviving patients in the data from the PREdiction of Clinical Outcomes from Genomic Profiles (PRECOG) study (Fig. 5B) (25). In two studies of melanoma patients treated with anti-PD-1 antibody after 28 days and 11 days, respectively (32,33), and a study of melanoma patients treated with combination anti-PD-1 plus anti-CTLA-4 antibodies (33), responders demonstrated the INTIRE gene signature. Remarkably, the INTIRE gene signature also distinguished between melanomas that subsequently responded to treatment with anti-PD-1 antibody from those that did not (32,33) (Fig. 5A and C). Finally, melanomas in patients who were depleted of B cells by administration or anti-CD20 (34) demonstrated an attenuated INTIRE gene signature (Fig. 5A and D), exemplifying the integrated nature of this immune response.

**Figure 5.**
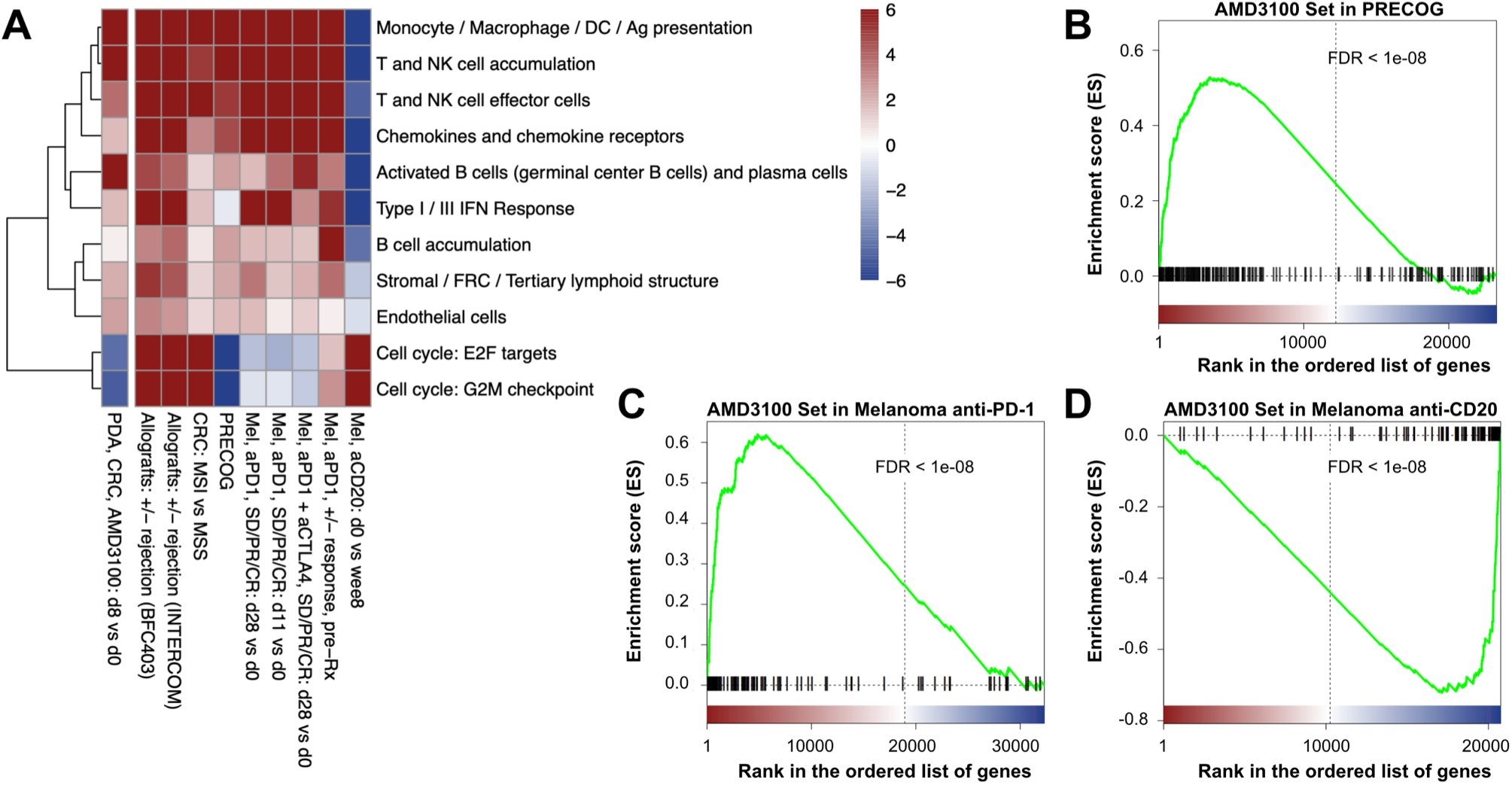
Comparative analyses of the integrated immune response (INTIRE) induced by AMD3100. (**A**) The heat map is shown of the enrichment analyses of nine gene sets representing different immune components that characterize the INTIRE in different immunological contexts, along with E2F target genes and genes involved in the G2M checkpoint. Comparative enrichment analyses for differentially expressed genes in paired biopsies with differential expressed genes from (**B**) biopsies from patients with longer versus shorter survival based on the PRECOG (25) analysis, (**C**) pre-treatment biopsies from patients with melanoma who responded to anti-PD1 treatment vs. non-responding patients (32,33), and (**D**) pre-treatment vs. week six of treatment biopsies from patients with melanoma who were depleted of B cell by treatment with anti-CD20 antibodies (34). (**A-D**) n=14 comprising of PDA (n=4) and CRC (n=10).

### Immune-mediated anti-cancer effects of AMD3100 administration

We examined the transcriptional changes in the paired biopsies for evidence of intratumoral immune mediated anti-cancer effects. Changes in the mRNA levels in biopsies from each patient of granzymes (GZM) A, B, H, K, and M and perforin, which encode the proteins that mediate killing by effector CD8^+^ T cells, significantly inversely correlated with changes in the mRNA levels of three genes uniquely expressed by cancer cells, CEACAM 5, 6, and 7 (Fig. 6A), but not with non-cancer-specific genes (Fig. S9).

**Figure 6.**
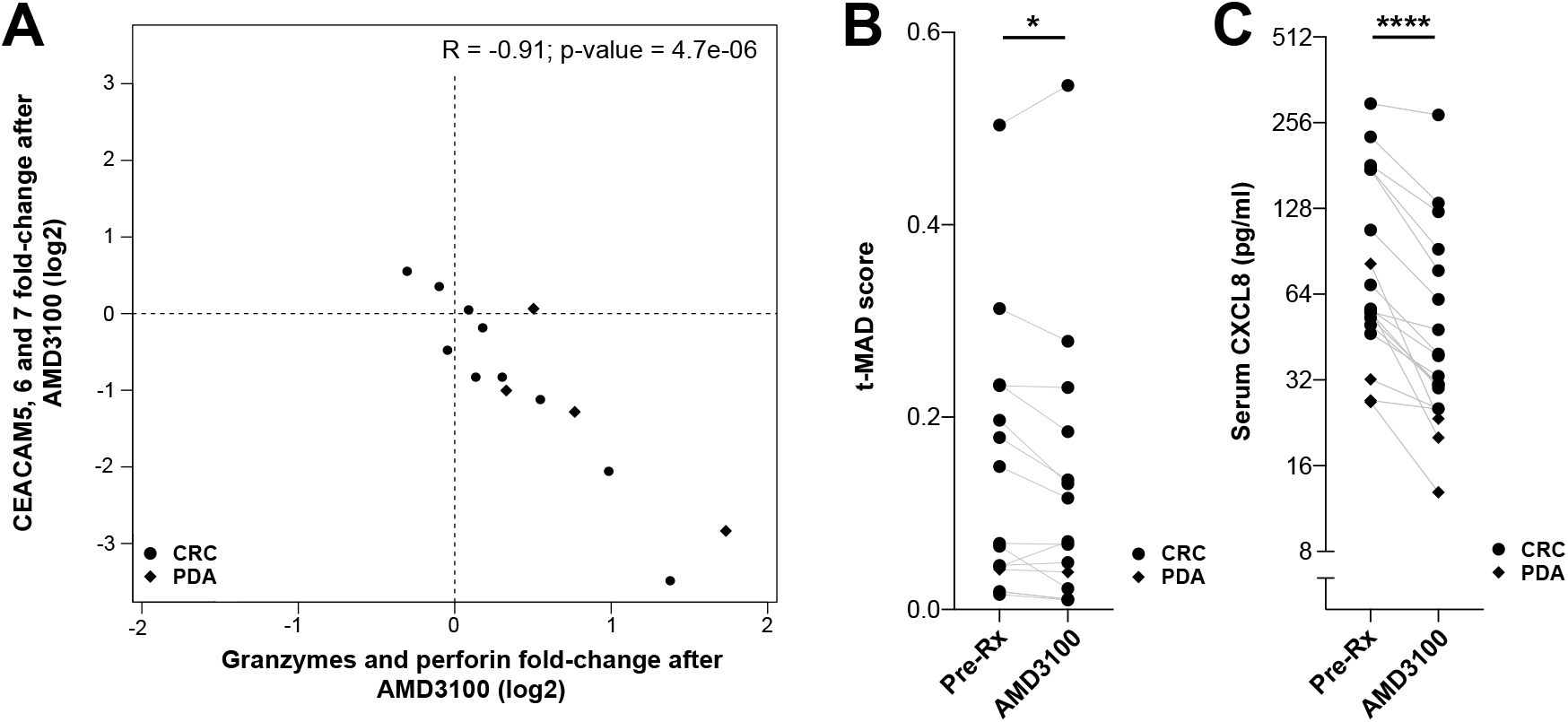
Analyses of the anti-cancer effect induced by AMD3100 treatment. (**A**) Changes of mRNA expression in the paired biopsies obtained from each patient with CRC (n=10) and with PDA (n=4) before and after treatment with AMD3100 for granzymes A, B, H, K, and M and perforin negatively correlate with changes in the expression of CEACAM 5,6, and 7. (**B**) Plasma ctDNA levels (n=15) and (**C**) serum concentrations of CXCL8 (n=18) pre-treatment (pre-Rx) and after seven days of continuous infusion of AMD3100 are shown. Statistical comparisons by Spearman’s rank correlation test (**A**), by paired Wilcoxon signed-rank test paired (**B**), and by paired t-test (**C**): *, p<0.05; ****, p<0.0001.

Next we evaluated plasma biomarker evidence of anti-cancer effects in all samples that passed the respective quality thresholds for analysis. The plasma concentrations of the tumor-derived markers, carcinoembryonic antigen (CEA) and carbohydrate antigen 19-9 (Ca19-9), which are not validated as early response markers in immunotherapy trials, were not significantly changed over the 7 day treatment period of the patients with AMD3100 (n=15, p=0.4). Next, we quantified circulating tumour DNA (ctDNA). Levels of ctDNA have been shown to decrease when patients respond to therapy (35–38). We evaluated ctDNA levels at baseline and after 7 days of treatment with AMD3100 (Fig. 6B). ctDNA levels were significantly reduced following treatment with AMD3100 (n=15, p=0.033). Furthermore, plasma levels of CXCL8, which has also been identified as a marker of tumour burden (39) and may provide an early indicator of therapeutic response (39–41), were also significantly decreased following treatment with AMD3100 (Fig. 6C, n=18, p<0.0001). The decrease in both ctDNA and CXCL8 levels support the possibility of an early anti-cancer effect mediated by CXCR4 inhibition.

## Discussion

The findings that the CAF mediates intratumoral immune suppression (18,20) and that a CAF-derived chemokine, CXCL12, coats the cancer cells in PDA and CRC suggest that its receptor, CXCR4, has a role in mediating immune suppression in the tumor microenvironment. The association of CXCL12 with cancer cells is predicted to have two immunological consequences. First, most immune cells in PDA and CRC tumors express CXCR4 and will, therefore, be stimulated by cancer cell-associated CXCL12 via ligation of this receptor. Second, CXCL12-stimulated CXCR4 inhibits the chemotactic functions of the chemokine receptors that direct the migration of immune cells. Therefore, the CXCL12-coat of cancer cells could impair the intra-tumoral accumulation of multiple immune cell types. We tested these two predictions in an experimental medicine study in which patients with PDA and CRC received the, small molecule CXCR4 antagonist, AMD3100, which is licensed for mobilization of hematopoietic stem cells. Treatment of patients with AMD3100 was limited to continuous iv infusion of the drug for one-week, using a protocol shown to be safe in patients with HIV (23). We first confirmed that the AMD3100 infusion did achieve continuous CXCR4 inhibition by observing the presence of CD34^+^ HSCs in the peripheral blood of each patient. All patients from the dose escalation phase showed persistent elevations of CD34^+^ HSCs, indicating the occurrence of continuous CXCR4 inhibition, and we therefore included patients from the entire cohort in all subsequent analyses. The short duration of drug administration limited our ability to observe whether CXCR4 inhibition induced clinical responses in patients with PDA or CRC, as assessed by standard radiological evaluations (42,43). These assessments did not reveal remissions and the lack of change in tumor volume is not informative due to the short time period that elapsed between scans. An independent clinical trial testing discontinuous CXCR4 inhibition by subcutaneous administration of a cyclic peptide inhibitor of CXCR4 together with anti-PD-1 antibody over several cycles in patients with advanced pancreatic cancer showed some evidence of clinical responses (44). We observed significant decreases in the levels of ctDNA and circulating CXCL8 of patients after treatment with AMD3100. ctDNA and CXCL8 are increasingly recognized as markers of tumour burden (35,38,39) and may provide early indications of response to therapy (35–41), when imaging evaluation is not conclusive (42,43). However, these intial observations will require further prospective validation.

This study focused on the question of whether the CXCL12-coating of cancer cells in PDA and CRC signified the existence of a fundamental immune suppressive pathway in two human cancers that have thus far resisted cancer immunotherapy. We chose to detect immunological changes by performing bulk RNASeq analysis of paired biopsies of metastatic lesions taken from patients before and at the end of the AMD3100 infusion. This analysis provided an unbiased and quantitative means of measuring the AMD3100-induced changes in the complex intratumoral immune environments of these tumors, which could be compared to similar transcriptional analyses of tissues representing other immunological reactions.

This comparative transcriptional analysis revealed unanticipated similarities between the immunological effects of two mechanistically distinct immunotherapies, inhibition of T cell checkpoints and inhibition of a chemokine receptor, respectively, in cancers that have different developmental origins, adenocarcinomas and melanomas. Both anti-PD-1 and anti-CTLA-4 antibody therapies, which enhance the activation of T cells, and AMD3100 treatment, which affects the trafficking of immune cells, up-regulated the expression of genes that characterize rejecting renal allografts, an example of immune damage to immunogenic, non-cancer tissue. Thus, effective cancer immunotherapy engages an immune pathway that mediates damage to non-infected, immunogenic tissue. This pathway involves multiple immune elements, and their participation could be assessed by the INTIRE signature, which characterizes nine components of the immune reaction. This analysis showed that the INTIRE signature was induced not only by CXCR4 inhibition in patients with PDA and CRC, but also by successful treatment of patients with melanoma with anti-PD-1 antibody. The occurrence of the INTIRE signature was even predictive of subsequent clinical responses to anti-PD-1antibody therapy in patients with melanoma. The additional finding that AMD3100 leads to the increased frequency of FAP+ cells expressing CCL19, which is a characteristic of FRCs, is consistent with the concept that shifting the balance from immune suppressive fibroblastic cells to those with immune-enhancing functions improves the outcome of cancer immunotherapy. This observation is reminiscent of the recent reports that the presence of TLSs, which in the mouse requires FAP+ fibroblasts (15, 45), correlates with clinical responses to T cell checkpoint therapy (11–13).

Finally, the study raises the possibility that a substantial proportion of patients with MSS PDA and MSS CRC have on-going anti-cancer immune responses. A majority of the patients treated with AMD3100 showed enhanced intra-tumoral immune B and T cells responses after only seven days, which would be unusually rapid for a primary immune response. Thus, intra-tumoral immune suppression rather than immune ignorance may be a major barrier to clinically effective immunotherapy. This possibility should be assessed with an appropriate clinical trial of repeat cycles of continuous CXCR4 inhibition in combination with a T cell checkpoint antagonist.

## Data Availability

The data relating to the differential gene expression analysis are available as supplemental material.

## Acknowledgements

We thank all patients.

The study was carried out at/supported by the NIHR Cambridge Clinical Research Facility.

We acknowledge support from the Human Research Tissue Bank, supported by the NIHR Cambridge Biomedical Research Centre, for sample processing, and the Histopathology Core Facility at the CRUK CI for sample immunostaining and imaging.

We thank Purity Bundi, Breanna Demestichas, Nikos Demiris, Kate Donoghue, Alex Overhill, Richard Houghton, and Eva Serrao for help with data acquisition, trial management, and data illustration and Hannah Meyer for critical appraisal of the manuscript.

## Funding

SU2C-Lustgarten Foundation Dream Team. Cancer Research UK Institute core grants C14303/A17197 and C9545/A29580. The Li Ka Shing Centre in which some of this research was performed was generously funded by CK Hutchison Holdings Limited, the University of Cambridge, Cancer Research UK, The Atlantic Philanthropies and a range of other donors.

Individual funding acknowledgements:

DIJ, FR: CRUK C14303/A17197 and C9545/A29580

TJ: CRUK C42738/A24868; National Institutes of Health USA 5P30CA045508-31; Pershing Square Innovation Fund.

CMC: Experimental Medicine Initiative Clinical Lectureship

## Conflict of Interest

Sanofi provided study drug for the clinical trial and validation of the PK assay, but had no part in study design, data acquisition, data analysis, or manuscript preparation.

## Current employment

MS: Odette Cancer Center, Sunnybrook Health Sciences Centre, Toronto, Ontario, Canada LM: AstraZeneca, R&D Oncology, The Anne McLaren Offices, C/O Academy House, Central Cambridge, 136 Hills Road, Cambridge CB2 0QH.

## Methods

### Immunofluorescence of human tumor arrays

Formalin fixed, paraffin embedded (FFPE) human pancreatic and colorectal tumor arrays (US Biomax) were deparaffinized in Xylene, washed three times with ethanol, and rehydrated in a serial concentration of ethanol (from 95% to 50%) and finally in water. For antigen retrieval, the sections were boiled in 10 mM Tris, pH 8.8 plus 1 mM EDTA for 10 minutes followed by cooling down for 30 minutes, a wash with PBS, and blocking with 1% BSA/PBS at room temperature for 1 hour. Following two washes with 0.05% Tween-20/PBS and one with PBS, Alexa Fluor 568 conjugated anti-KRT19 antibody (Abcam, ab203445) and FITC conjugated anti-CXCL12 antibody (R & D Systems, IC350F) were applied and the sections were incubated at room temperature for 1 hour. Finally, the sections were stained with DAPI (Thermo, R37606) for 10 minutes and washed with 0.05% Tween-20/PBS for two times and once with PBS, followed by application of mounting medium (Thermo, P36961) and imaging with Leica SP8 confocal microscope. Images were analyzed and exported through ImageJ.

### Chemotaxis assays

#### Plasmids

To generate lentiviral plasmid expressing human chemokine receptors, CXCR1 and CXCR3 cDNA from CXCR1-Tango (Addgene, #66259) and CXCR3-Tango (Addgene, #66261), respectively, were amplified and subcloned into lentiCas9-blast (Addgene, #52962) with restriction enzymes Age1 and BamH1 to replace SpCas9; CXCR5 and CXCR6 cDNA from CXCR5-Tango (Addgene, #66263) and CXCR6-Tango (Addgene, #66264), respectively, were amplified and subcloned into lentiCas9-puro to replace SpCas9 where blasticidin resistant gene (blast) was also replaced with puromycin resistant gene (puro). For CRISPR editing, control guide (sgScramble, GCTTAGTTACGCGTGGACGA) and guides targeting to human *CXCR4* gene (sgCXCR4-1, TGACATGGACTGCCTTGCAT; sgCXCR4-2, CAACCACCCACAAGTCATTG; sgCXCR4-3, CAGGACAGGATGACAATACC) or human *RGS14* gene (sgRGS14-1, GCAGGGATCTGTGAGAAACG; sgRGS14-2, TCGGCAGCCCTGACGCCACG; sgRGS14-3, CTGAGACTCTCGGCGCAAGG) were cloned into the vector lentiCRISPR-v2 (Addgene, #62961).

#### Cell lines

Jurkat cells (ATCC, Clone E6-1, TIB-152) were transduced with lentivirus expressing human CXCR1, CXCR3 or CXCR6, followed by treatment with 5 µg/ml blasticidin (CXCR1 and CXCR3) or 0.5 µg/ml puromycin (CXCR6) for two weeks, to generate Jurkat-CXCR4/CXCR1 cells, Jurkat-CXCR4/CXCR3 cells and Jurkat-CXCR4/CXCR6 cells, respectively. For Raji B-CXCR4/CXCR5 cells, Raji B cells (ATCC, CCL-86) were transduced with CXCR5 lentivirus followed by selection with 0.5 µg/ml puromycin for two weeks. Sub-population of CCRF-HSB-2 cells (ATCC, CCL 120.1) that spontaneously express high level of CXCR3 were FACS sorted as HSB2DP-CXCR4/CXCR3 cells, where DP stands for “double positive”. The Molm13 cell line (Molm13-CXCR4/CCR2) which expresses both CXCR4 and CCR2 spontaneously was a gift from Dr. Vakoc’s laboratory. For CRISPR knock-outs, Jukat-CXCR4/CXCR3 and Molm13-CXCR4/CCR2 cells were transduced with lentivirus expressing SpCas9 and control guide (sgScramble), guides targeting to CXCR4 (sgCXCR4) or RGS14 (sgRGS14). HSB2DP-CXCR4/CXCR3 cells were cultured in IMDM medium (ATCC, 30-2005) supplemented with 10% FBS (Seradigm, 1500-500), 100 units/ml penicillin and 100 µg/ml streptomycin. All the other cell lines were cultured in RPMI-1640 medium (ATCC, 30-2001) plus 10% FBS and penicillin/streptomycin. Cells were maintained at the density of 1⨯10^5^-1⨯10^6^ cells/ml.

#### Flow cytometry

For each cell line, 1⨯10^5^-1⨯10^6^ cells were pellet and washed with cold FACS buffer (PBS, 2% FBS and 20 mM HEPES, pH 7.4), followed by Fc receptor blocking with Human TruStain FcX (BioLegend, 422301) in the FACS buffer at 4°C for 30 min. Cells were then stained with APC conjugated anti-Human CXCR4 antibody (BioLegend, 306510) and/or Alexa Fluor 488 conjugated anti-Human CXCR1 antibody (BioLegend, 320616), PE conjugated anti-Human CXCR3 antibody (BioLegend, 353706), FITC conjugated anti-Human CCR2 antibody (BioLegend, 357215), FITC conjugated anti-Human CXCR5 antibody (BioLegend, 356913), APC conjugated anti-Human CXCR6 antibody (R & D Systems, FAB699A) for another 30 min at 4°C. Then, cells were washed with the FACS buffer twice and analyzed with BD LSRForsseta cell analyzer.

#### Chemotaxis assays

The chemotaxis buffer for HSB2DP-CXCR4/CXCR3 cells is IMDM medium plus 0.1% BSA and 20 mM HEPES, pH 7.5 and for all other cell lines is RPMI-1640 medium plus 0.1% BSA and 20 mM HEPES, pH 7.5. After collection, cells were washed for two times and re-suspended in the chemotaxis buffer at the density of 1⨯10^6^/ml, followed by incubation at 37°C for 30 min. For the CXCL12 cross-inhibition of the other chemokine induced chemotaxis, 80 µl cells with or without 100 ng/ml (200 ng/ml for CCR2) recombinant human CXCL12 (R & D Systems, 350-NS) were loaded in the upper wells of the Boyden chamber plate (VWR, 89089-934; Supplier NO. 3388, 5 µm membrane pore size), and the lower chambers were flowed in 240 µl chemotaxis buffer with PBS or the other specific recombinant human chemokine. To test the effect of other specific chemokine on the CXCL12 induced chemotaxis, cells and the specific chemokine were loaded on the upper chamber, with CXCL12 containing chemotaxis buffer loaded in the lower chamber. To evaluate the activity of AMD3100 (Sigma, A5602) on CXCL12 or the other specific chemokine induced chemotaxis, cells were pre-treated with increasing dose of the drug at 37°C for 30 min, then the cell/AMD3100 mixture was loaded in the upper chambers, and CXCL12 or other specific chemokine containing chemotaxis buffer was loaded in the lower chambers. For the AMD3100 rescue of the CXCL12 mediated cross-inhibition, the AMD3100 pre-treated cells together with 100 ng/ml CXCL12 were loaded in the upper chambers, and chemotaxis buffer containing the other specific chemokine was flowed in the lower chambers. After the experiments were set up, the Boyden chamber plates were incubated at 37°C for 2 or 3 hours. Then, the number of cells in the lower chamber was counted with a Guava bench-top flow cytometer, for 30 seconds at medium flow rate (∼1.2 µl/s). The concentration of each chemokine used is as following: CXCL8 (R & D Systems, 208-IL), 20 ng/ml; CXCL10 (R & D Systems, 266-IP), 1000 ng/ml; CXCL13 (R & D Systems, 801-CX), 1000 ng/ml; CXCL16 (R & D Systems, 976-CX), 50 ng/ml; CCL2 (R & D Systems, 279-MC), 200 ng/ml.

### RNA sequencing analysis

#### Gene expression quantification and differential gene expression analysis

Sequences of human transcripts were downloaded from Ensembl release 97. Transcripts quantification was performed using Kallisto ver. 0.43 and gene-level count matrices for use in DESeq2 were calculated using *tximport* as recommended by DESeq2 authors (46). All subsequent analyses on gene expression were performed using R 3.5.0. For differential expression analysis, raw counts were used directly in DESeq2. For other downstream analyses (i.e. fold-change correlation plots) counts data were transformed using the variance stabilizing transformation (VST) as implemented in DESeq2.

#### Gene set enrichment analysis (GSEA)

Gene set enrichment analysis was performed using the *fgsea* package available in Bioconductor. GSEA plots were obtained using a modified version of the associated *plotEnrichment* function. All gene sets used in the manuscript are provided as supplementary table. Data used for Figure 5A was obtained from several different sources: the rejecting allografts gene list was obtained from GEO series GSE48581 (29) and GSE36059 (28); the microsatellite instability gene list was obtained by combining Xena UCSC TCGA gene expression data (31) and MSI status for the same tumor samples (30); the list of genes associated with pan-cancer survival was obtained from the PRECOG portal (25); the anti-PD1 response gene lists were obtained from GEO: GSE91061 (32) and ENA: PRJEB23709 (33); the anti-CD20 gene list was obtained from Array Express: E-MTAB-7473 (34).

#### Code availability

The authors declare that the R code used to generate the analysis presented in this study is available upon reasonable request.

### Experimental medicine study

#### Patient eligibility

Patients with advanced or metastatic pancreatic adenocarcinoma (PDA), high grade serous ovarian cancer (HGSOC) or colorectal adenocarcinoma (CRC), refractory to or declining conventional chemotherapy were eligible for the dose escalation phase. The 10-patient expansion cohort at the recommended phase 2 dose (RP2D) of 80 µg/kg/hr was restricted to patients with PDA. Other eligibility criteria included a lesion accessible to biopsy, Eastern Cooperative Oncology Group (ECOG) performance status of 0 or 1, and adequate organ function including a lymphocyte count above the lower limit of normal. Patients were excluded if they had significant cardiac co-morbidities, such as past history of significant rhythm disturbance. Full eligibility criteria can be viewed at https://clinicaltrials.gov/ct2/show/NCT02179970. Patients were accrued at Cambridge University Hospitals NHS Foundation Trust and Weill Cornell Medicine/New York Presbyterian Hospital, NY, USA.

#### Study design

This phase 1, multicentre, open-label, non-randomized study used a 3+3 dose escalation design. The primary endpoint was the safety (evaluated using the Common Terminology Criteria for Adverse Events (CTCAE) 4.03) of AMD3100 administered, to achieve a plasma AMD3100 concentration at steady state ≥2µg/ml in ≥ 80% of patients at the RP2D. AMD3100 was administered as a 7-day continuous infusion. Dose-limiting toxicity (DLT) was defined as an adverse reaction (AR) ≥G3 occurring within 21 days of AMD infusion. Secondary endpoints included overall response rate (RECIST 1.1) at 14 (+/-2) days after the infusion, and metabolic changes in tumor using [^18^F]FDG-PET/CT within 1 day of infusion completion. Baseline scans were performed within 14 days before the start of the infusion. Exploratory objectives included the assessment of immune changes in tumor biopsies. Patients were monitored by cardiac telemetry for the initial 48hr of the infusion (later amended to 72hr), followed by Holter monitoring for the remainder of the infusion.

Patients provided written informed consent to Research Ethics Committee-approved protocol (REC reference 15/EE/0014 at the UK center and IRB number 1508016466 at the US center), in compliance with Good Clinical Practice (GCP), local regulatory requirements and legal requirements. A Clinical Trial Authorisation (CTA) was obtained from the Medicine and Healthcare Regulatory Authority (MHRA). The study was sponsored by Cambridge University Hospitals NHS Foundation Trust and the University of Cambridge and Weill Cornell Medicine/New York Presbyterian Hospital.

#### AMD3100 sample and pharmacokinetic analysis

AMD3100 plasma concentration was assessed at the following nominal time points: pre-dose, 24, 72 and 168 hrs of the infusion. A time point at day 7 (+/-2) after infusion discontinuation was added from patient 1017 onwards. The concentration data were generated using a liquid chromatography-tandem mass spectrometry (LC-MS/MS) method, that met the requirements of the EMA guidance on method validation, performed by the CRUK Cambridge Institute PK/Bioanalytics Core Facility. A claim of GCP compliance is made for the sample analysis data, pending the demonstration of long-term storage stability, which is on-going at the time of publication. AMD3100 calibration standards were prepared in the range of 40-4,000 ng/mL using blank control human plasma obtained from the NHS blood transfusion service (lower limit of quantification (LLOQ) 40 ng/mL). After the addition of the internal standard (AMD3100-D4) and EDTA (10mM), plasma samples, quality control (QC) and calibration standards were extracted by protein precipitation with 1% formic acid in methanol. A portion of the supernatant was evaporated to dryness and the residue reconstituted in 1% formic acid in water prior to analysis on the LC-MS/MS. High-performance liquid chromatography was performed with the Shimadzu Nexera X2 using a Phenomenex Kinetex F5 column (1.7µm, 100 ⨯ 2.1mm) and mobile phases A and B containing 0.1% formic acid in water or methanol, respectively. MS/MS detection was carried out using a Sciex API6500 mass spectrometer with an electrospray source. QC samples (120, 400 and 3000 ng/mL) were used to determine the precision (coefficient of variation [%CV]) and accuracy (relative error [%RE]). QC intra-day %CV was ≤9.8%, and %RE ranged between −3.1 to 4.8%. All instrument control and data collection was performed using Analyst v1.6.2, and peak area integration, regression and quantification using MultiQuant(tm) v3.0.2. A weighted (1/x2) least square linear regression was used to construct the calibration line.

### Pharmacodynamic analysis

At baseline, 24/72/168hr of the infusion, and 21(+/-2) days after infusion, blood was collected into Sarstedt EDTA blood tubes and CD34+ cells quantified by flow cytometry. The CD34+ absolute count was derived from a bead-based assay using BD Bioscience Trucount tubes and an ISHAGE gating strategy.

### Positron Emission Tomography

Positron Emission Tomography (PET) was performed in conjunction with low dose Computed Tomography (CT) for attenuation correction and localization purposes. Images were acquired from the midbrain to the knees, approximately 90 (89.7 ± 21.1) min after the injection of ∼370 (368 ± 13) MBq of [18F]fluorodeoxyglucose ([18F]FDG). The baseline [18F]FDG-PET/CT was performed before or at least 24 hours after the core tissue biopsy, and the post treatment PET/CT was performed within 24 hours after the biopsy on day 8 of the study. 19 patients underwent imaging at both timepoints and were evaluated for metabolic changes following treatment.

Response was determined by calculating the change in the mean weighted average of the Standardized Uptake Value (SUVMWA) of the [18F]FDG avid target lesions. SUVMWA was calculated as the sum of the product of the SUVmean75% and volume for all target lesions, divided by the total target lesion volume. A clinically significant change was defined as change in SUVMWA ≥ 30%.

### Biopsy processing

#### RNA and DNA extraction

Snap frozen biopsies were embedded in chilled OCT (-4 °C) and allowed to solidify in a cryotome at −20 °C. Subsequently, ten 30µm sections were cut and collected in RLT buffer for DNA/RNA extraction, followed by six 6µm sections for histologic analysis. The remaining tissue was cut in 30µm sections until exhausted. All 30µm sections were extracted for DNA and RNA using the AllPrep DNA/RNA/miRNA Universal Kit (Qiagen, 80224) according to manufacturer instructions from tissues, and DNA quantification by fluorometer according to manufacturer instructions (Qubit 3.0, Life technologies). An H&E was reviewed by a Histopathologist to determine cellular content (> 40% cancer cell content).

#### Histopathologic analyses

3 µm formalin-fixed, paraffin-embedded (FFPE) tissue sections were deparaffinised in xylene and rehydrated in an ethanol series. Immunofluorescence staining was performed on the Bond Rx automated platform. Sections were retrieved with Tris EDTA for 20 minutes and incubated sequentially with primary and secondary antibody pairs (where applicable) at room temperature for 30 minutes or, for CXCL12, 60 minutes. Slides were counterstained with DAPI and digitalized using the Axio Scan.Z1 (Zeiss) at 20 ⨯ (0.22µm/ pixel). Tumour areas were selected based on review of serial H&E sections by a histopathologist: liver, peritoneal fat and necrosis were excluded. The HighPlex FL v3.0.1 algorithm in Halo software (v2.3, Indica labs) was used to automatically detect CD8 positive cells across all sections, with a tissue classifier that restricted analysis to pan-CK positive areas within the predefined tumour area. The same algorithm was used for pre and post treatment biopsies. Cell counts were normalised to pan-CK tissue area (cells µm^-2^).

Primary antibodies: CD8 (SP16; Lab Vision/Thermo Scientific), pan-cytokeratin (AE1/ AE3 Alexa Fluor 488 conjugate; eBioscience), CXCL12 (79018; R&D).

### Immunological assays

#### Fluorescent in-situ hybridization

RNAscope (Advanced Cell Diagnostics, ACD) was performed following the standard protocol. Frozen sections were fixed with 4% paraformaldehyde (PFA) for 15 min at 4°C and dehydrated with sequential ethanol solution (50%-100%) at room temperature. Sections were incubated with pretreat IV for 30 min and washed in phosphate-buffered saline before being hybridized with gene-specific probes (CCL19, FAP) for 2 h at 40°C in a HybEZ oven. Sections were washed with wash buffer followed by incubations in sequential amplifers (Amp1-Amp4). Finally, sections were stained with DAPI and mounted with prolong antifade mountant. Images were taken by Leica SP8 confocal microscope and analyzed with ImageJ software.

### Plasma cell-free DNA analysis

Blood samples were collected into EDTA-containing tubes and processed by a double-centrifugation protocol (1,600 g for 10 minutes; 14,000 rpm for 10 minutes) before storage at − 80°C. Plasma DNA was extracted using QIAsymphony DSP Circulating DNA Kit (QIAGEN) and DNA sequencing libraries were prepared using the ThruPLEX Plasma-seq kit (Takara Bio). Unique DNA barcode sequences were introduced to allow pooled sequencing runs on HiSeq 4000 (Illumina) generating 150-bp long paired-end reads. Sequencing reads were subsequently aligned to the human reference genome (hg19) using BWA-mem. PCR and optical duplicates were marked using MarkDuplicates (Picard Tools) and excluded from downstream analysis along with reads of low mapping quality. Sequencing data in each sample were downsampled to 5 million reads to generate t-MAD scores (47) for estimating ctDNA levels inferred from a genome-wide copy number aberration profile. The segmentation step was summarized by a median value and a t-MAD threshold of 0.015 was used.

### Cytokine analysis

Serum was analysed for CXCL8 using the MesoScale Discovery Human 10-plex ProInflammatory Panel 1 kit (V-PLEX K15049D-2), as per manufacturer’s instructions. Analyses were conducted at the Core biochemical assay laboratory (CBAL) at CUH, NHS Foundation Trust.

## Supplementary figure legends

**Figure S1.**
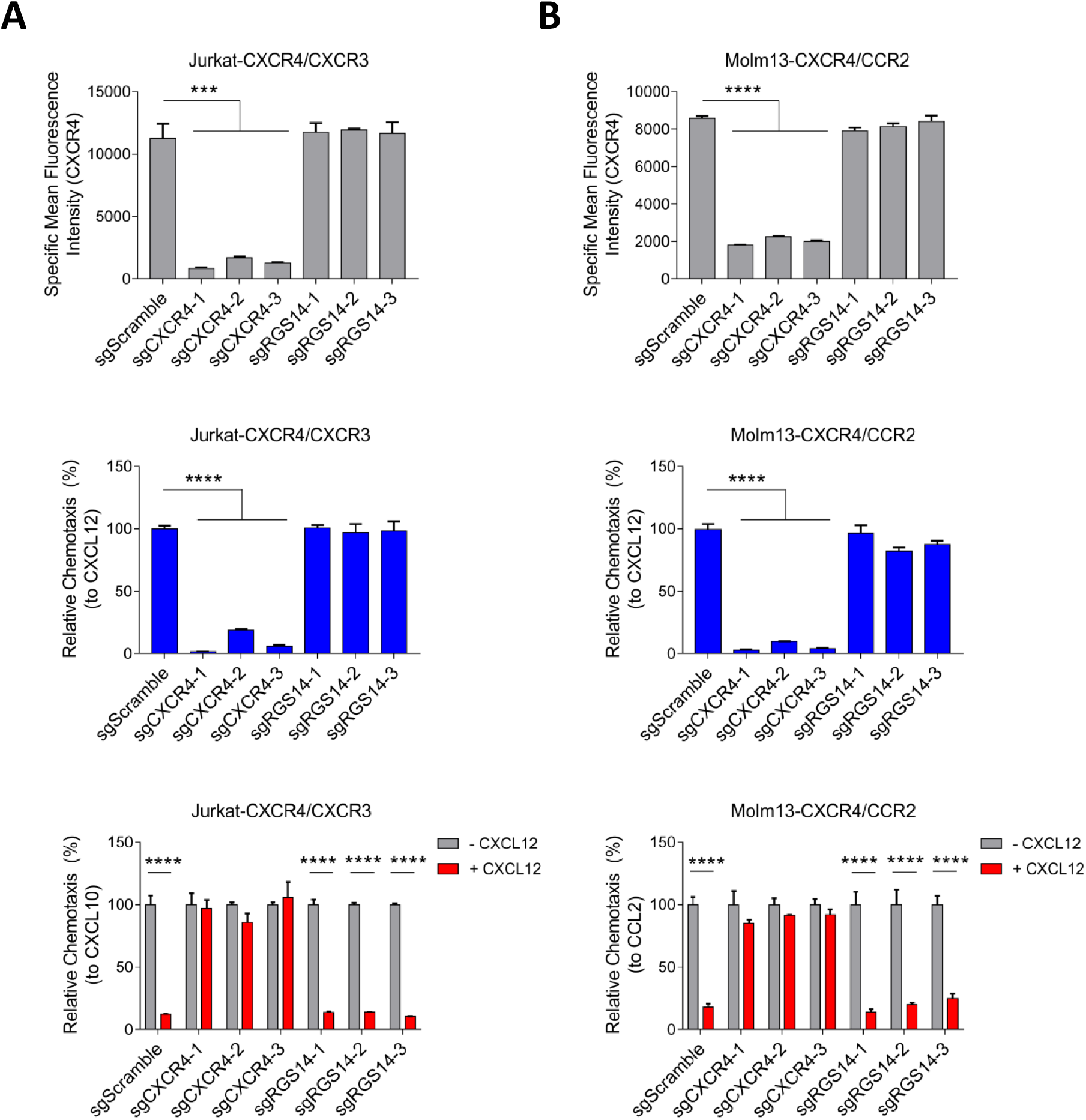
The inhibition by CXCL12 of CXCR3- and CCR2-mediated chemotaxis of human immune cells depends on CXCR4 co-expression. (**A**) Jurkat-CXCR4/CXCR3 T lymphoblastoid cells were CRISPR/Cas9-edited with different sgRNAs and assessed for CXCR4 expression by FACS analysis, chemotaxis to CXCL12 in the lower Boyden chamber, and chemotaxis to CXCL10 in the lower chamber in the presence of CXCL12 in the upper chamber. (**B**) Molm13-CXCR4/CCR2 monocytoid cells were CRISPR/Cas9-edited with different sgRNAs and assessed for CXCR4 expression by FACS analysis, chemotaxis to CXCL12 in the lower Boyden chamber, and chemotaxis to CCL2 in the lower chamber in the presence of CXCL12 in the upper chamber. Statistical analysis by student’s t test: ***, p<0.001; ****, p<0.0001.

**Figure S2.**
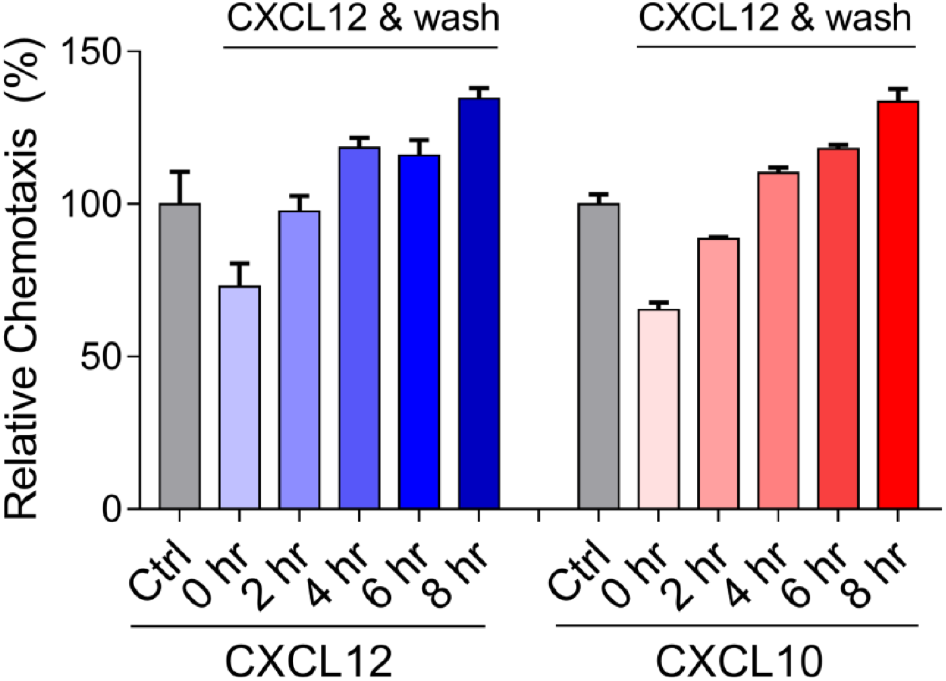
The reversibility of inhibition by CXCL12 of CXCL10-induced chemotaxis. HSB2DP-CXCR4/CXCR3 T lymphoblastoid cells were treated with PBS (Ctrl) or CXCL12 at 37°C for 15 minutes, followed by washing and resuspension in culture medium. After resting at 37°C for timed intervals, the cells were assessed for their chemotactic responses to CXCL12 (blue bars) or CXCL10 (red bars).

**Figure S3.**
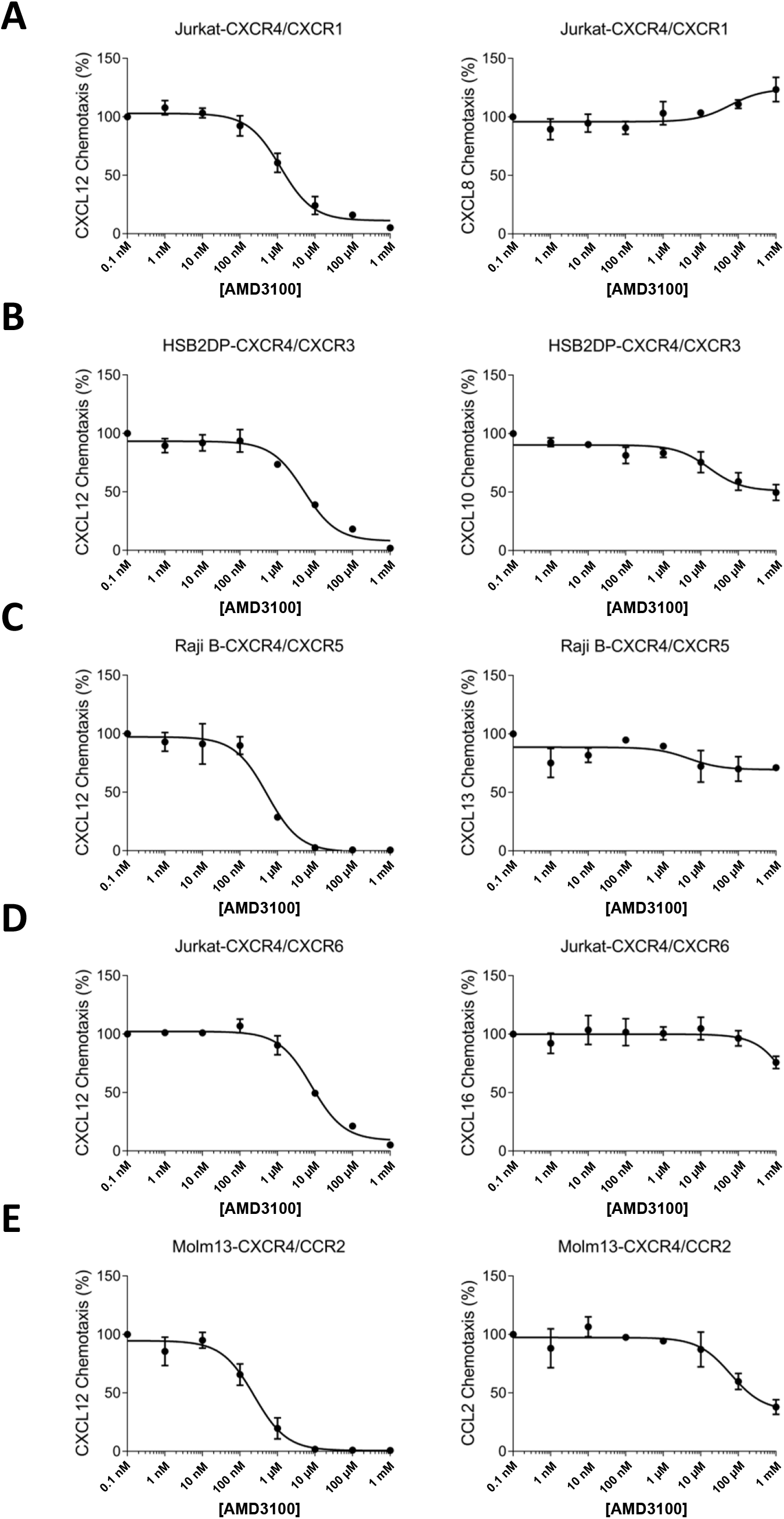
The effect of AMD3100 on the directed migration of the dual chemokine receptor-expressing human cells. The chemotactic responses of (**A**) Jurkat-CXCR4/CXCR1 cells, (**B**) HSB2DP-CXCR4/CXCR3 cells, (**C**) Raji B-CXCR4/CXCR5 cells, (**D**) Jurkat-CXCR4/CXCR6 cells, and (**E**) Molm13-CXCR4/CCR2 cells was assessed in the presence of the indicated chemokines and increasing concentrations of AMD3100. The results are expressed as percentage of the responses occurring in the absence of AMD3100.

**Figure S4.**
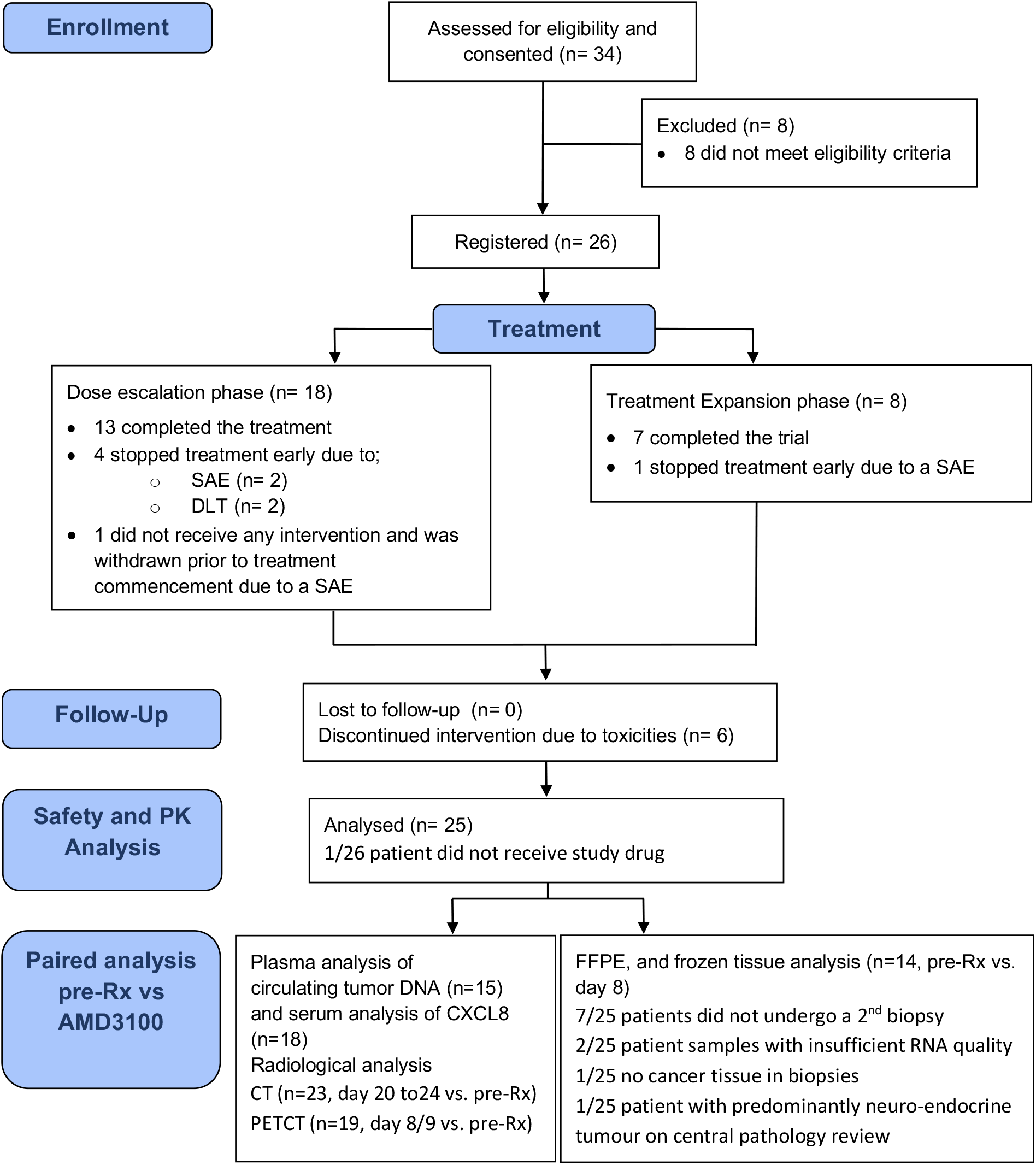
Consort diagram of the experimental medicine study (NCT02179970).

**Figure S5.**
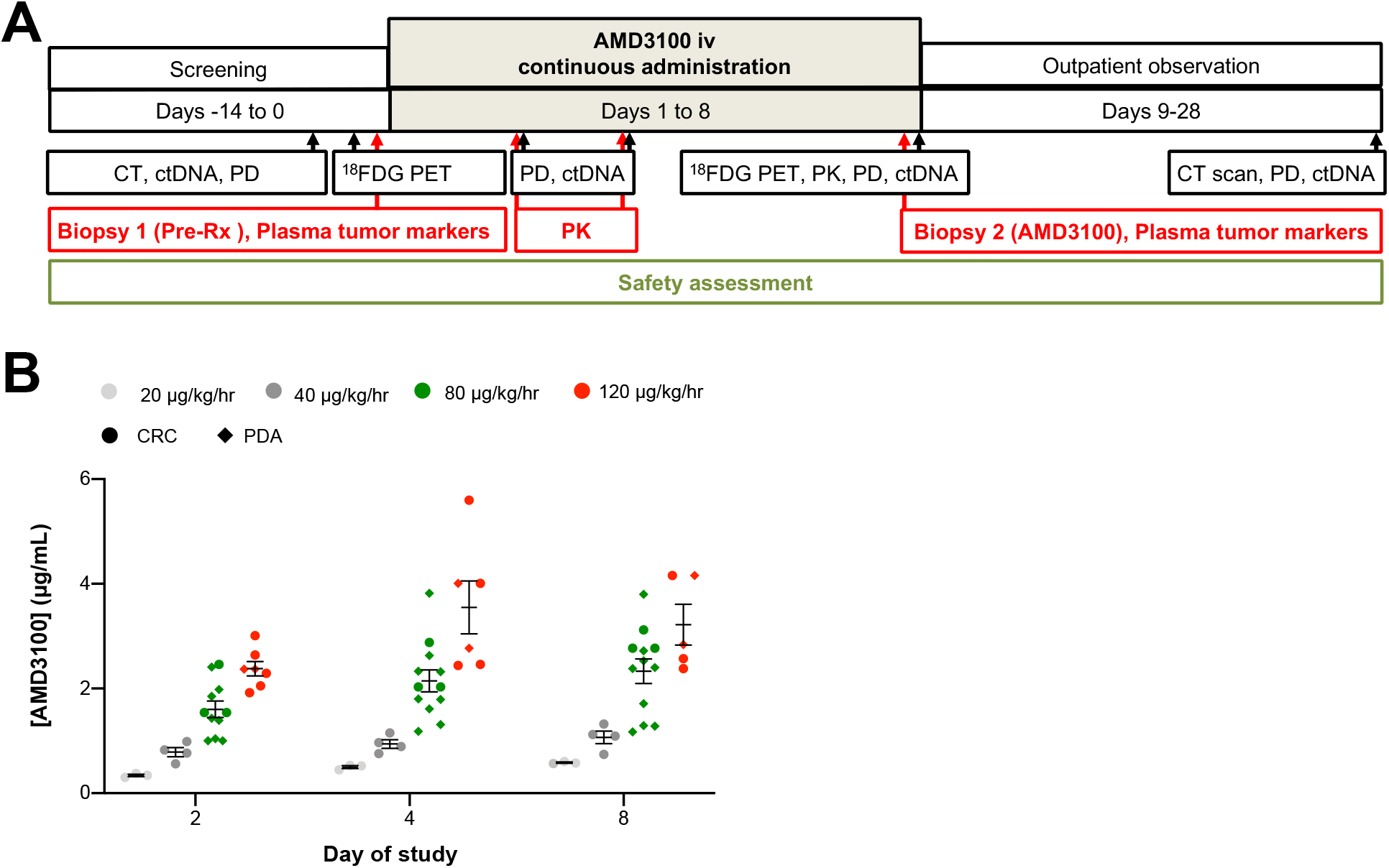
Schematic of the experimental medicine study and the pharmacokinetic and toxicity results. (**A**) The study time line is shown for the experimental medicine study of continuous iv infusion of AMD3100 to patients with PDA and CRC. (**B**) Plasma concentrations of AMD3100 at increasing dose levels of the iv infusion on day 2, 4, and 8 of the study are displayed. The mean and standard error of the mean are indicated.

**Figure S6.**
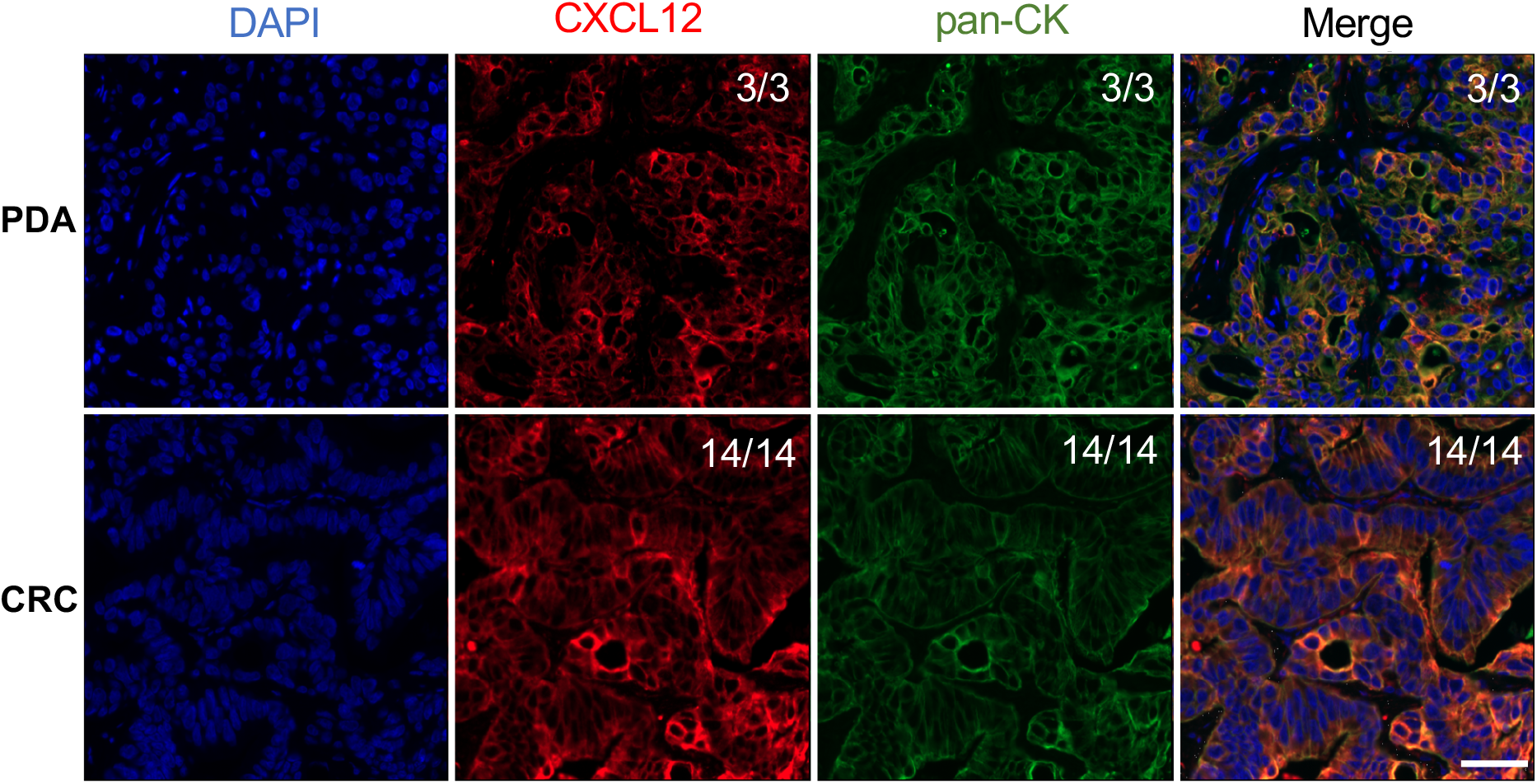
The CXCL12-coat on pancreatic and colorectal cancer cells from biopsies of study patients. For patients enrolled during the dose escalation part of the study, sections of biopsies of metastases from patients with pancreatic (PDA) and colorectal (CRC) adenocarcinoma were stained with fluorescent antibodies to pan-CK to reveal cancer cells, and to CXCL12. The ratios shown in the top right corners of the photomicrographs indicate the frequency of the observed staining relative to the total number of independent tumors assessed. Scale bar, 50 µm.

**Figure S7.**
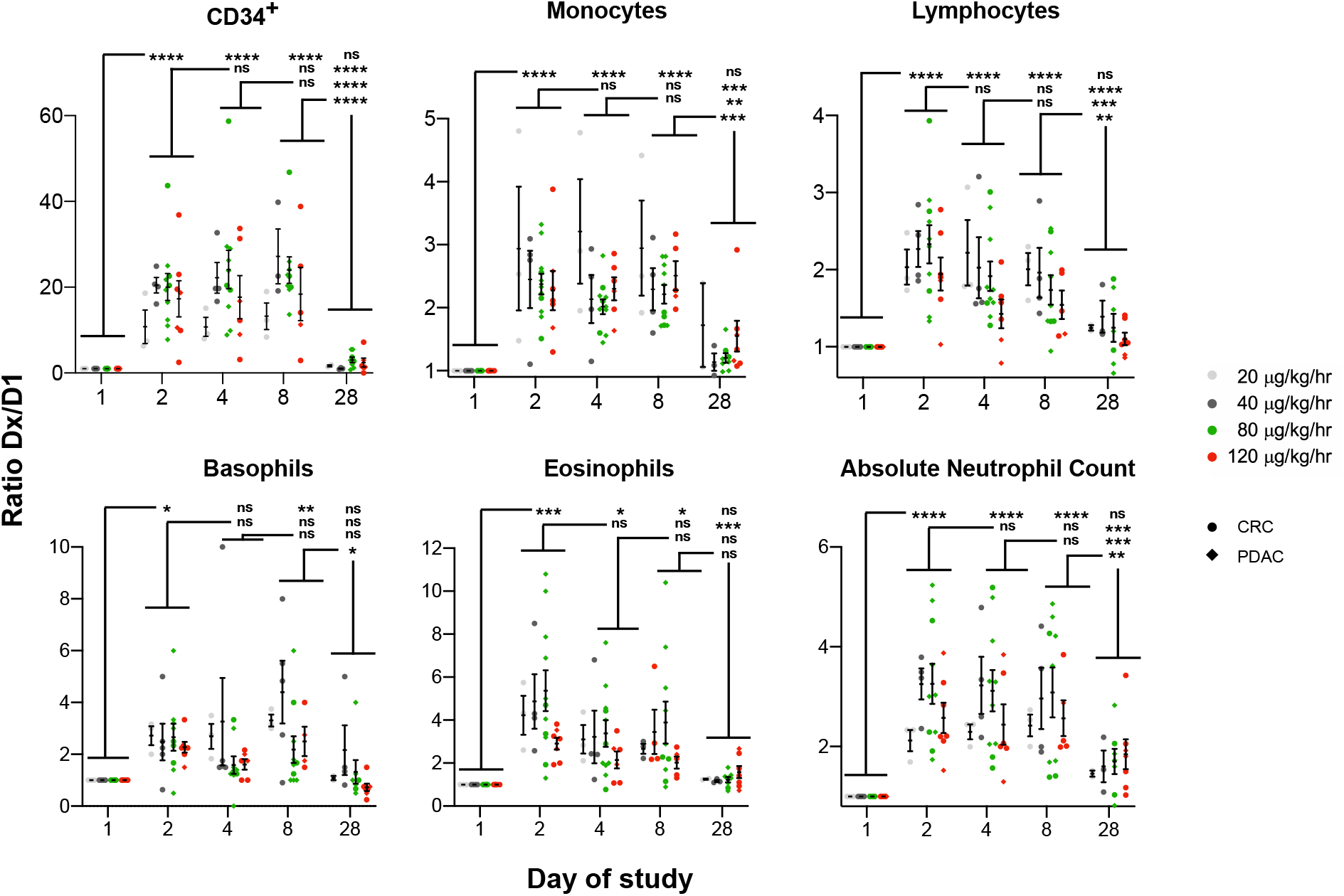
The effect of continuous AMD3100 administration on peripheral blood leukocyte levels. The relative levels for six leukocyte subtypes on study day two, four, eight, and 28, as compared to the levels of day one, are displayed. Subdivisions by dose level of AMD3100 are included for each study day. Statistical comparison of study day levels by nested ANOVA: *, p<0.05; **, p<0.01; ***, p<0.001; ****, p<0.0001; ns, not significant.

**Figure S8.**
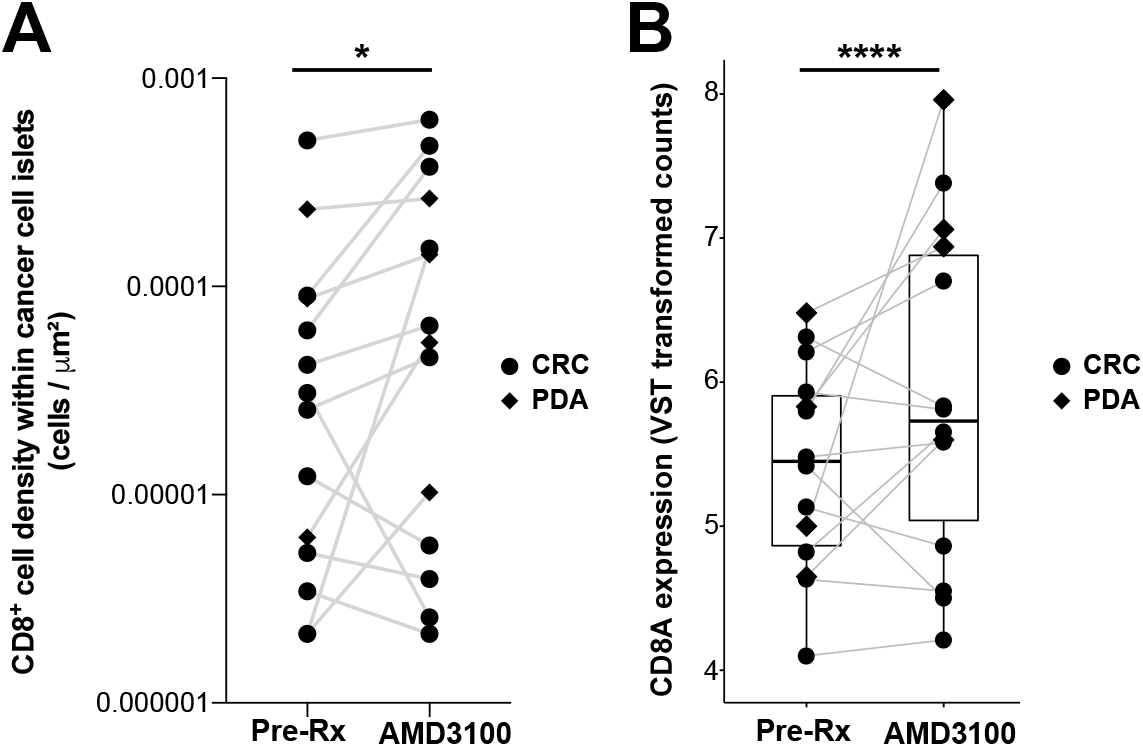
Quantification of immunehistochemical and transcriptional analyses of the frequency of intratumoral CD8^+^ T cells. Paired biopsies of metastases obtained from each patient with CRC or PDA were analyzed. (**A**) The frequency of CD8^+^ T cells, assessed by immunofluorescent staining of FFPE tissue, within cancer cell islets is displayed. (**B)** The mRNA levels of CD8A, assessed by RNAseq analysis, are shown. (**A** and **B**) n=14 comprising of PDA (n=4) and CRC (n=10). Statistical comparisons by paired t-test (**A**), and by Wald test (**B**): *, p<0.05; ****, p<0.0001.

**Figure S9.**
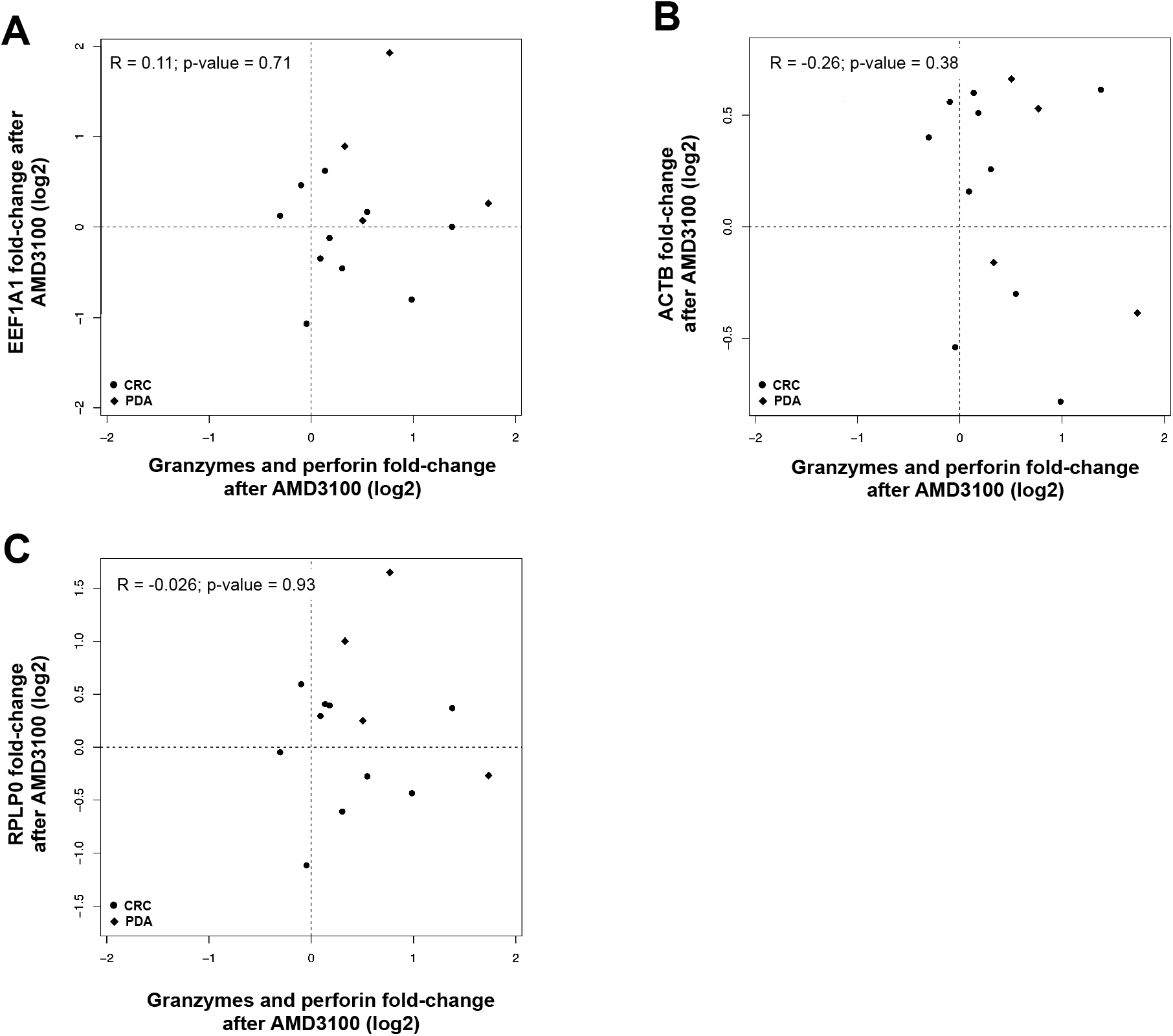
Correlation of granzymes and perforin with the expression of non-cancer-specific genes after AMD3100 treatment. Changes of mRNA expression after AMD3100 administration for granzymes A, B, H, K, and M and perforin compared to changes in mRNA expression for (**A**) EEF1A1, (**B**) ACTB, and (**C**) RPLP0 are shown. Statistical comparisons by Spearman’s rank correlation test are shown.

## Supplemental Tables and Figures

Biasci *et al*. CXCR4 inhibition in human pancreatic and colorectal cancers induces an integrated mmune response

**Table S1.** Early drug discontinuations due to adverse events.

| Table S1. Early drug discontinuations due to adverse events |  |  |  |  |  |  |
| --- | --- | --- | --- | --- | --- | --- |
| Dose cohort<br>(µg/kg/hr) | Patient<br>ID | Day of<br>discontinuation<br>or interruption | Adverse event | Relation to plerixafor | Action | Outcome of adverse<br>event |
| 40 | 1007 | 7 | Biliary tract obstruction and infection | Unlikely to be related | Discontinued | Recovered |
| 120 | 1013 | 5 | Duodenal obstruction | Unlikely to be related | Discontinued | Recovered |
| 120 | 1014 | 8 | Vasovagal reaction* | Possibly related | Discontinued | Recovered |
| 120 | 1018 | 2 | Vasovagal reaction, hypotension, abdominal pain* | Possibly related | Discontinued | Recovered |
| 80 | 1022 | 4 | Panic attack | Possibly related | Discontinued | Recovered |
| * Dose Limiting Toxicity |  |  |  |  |  |  |

**Table S2.**
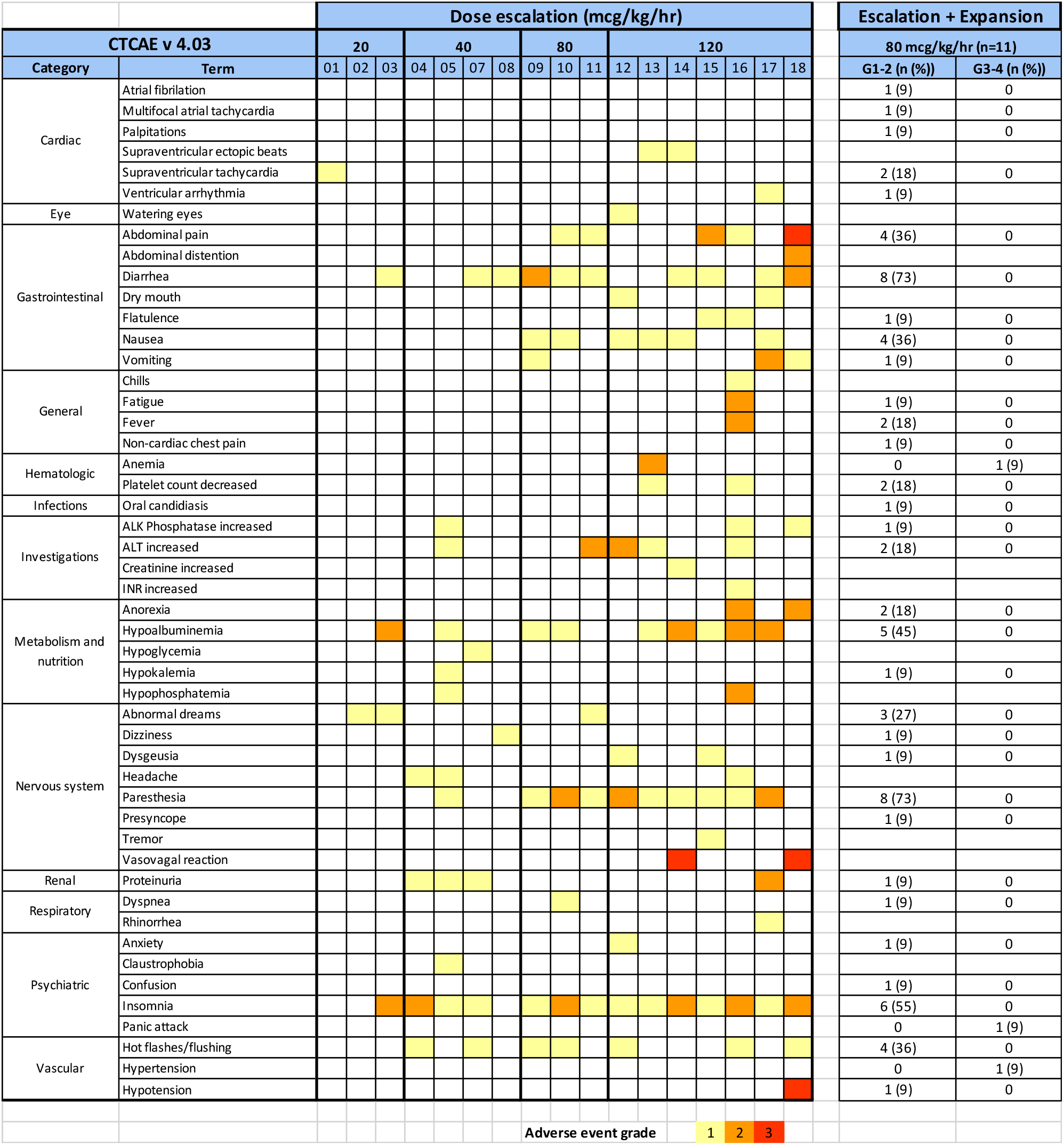
Drug related adverse events during dose escalation and expansion phase.

**Table S3.** AMD3100 pharmacokinetic parameter stimates.

| **Dose cohort**<br>**(µg/kg/hr)** | **Patients**<br>**enrolled** | **C<sub>ss</sub>**<br>**(µg/ml)** | **AUC<sub>0-168</sub>**<br>**(µg\*h/ml)** | **CI**<br>**(L/hr)** |
| --- | --- | --- | --- | --- |
| 20 | 3 | 0.58 (0.57-0.60) | 76 (72-80) | 2.86 (2.64-3.10) |
| 40 | 4* | 1.05 (0.74-1.32) | 143 (110-174) | 2.76 (2.67-2.92) |
| 80 | 11# | 2.28 (1.17-3.80) | 324 (182-544) | 2.82 (1.90-3.53) |
| 120 | 7† | 2.99 (2.38-4.16) | 431 (359-570) | 3.14 (2.45-3.74) |

Mean (range)
\* Patient 1007 withdrawn at day 6, excluded

## Notes

### Competing Interest Statement

The authors have declared no competing interest.

### Clinical Trial

NCT02179970

### Author Declarations

UK center, University of Cambridge: REC reference 15/EE/0014 US center, Weill Cornell Medicine: IRB number 1508016466

## References

1. Hodi FS, O’Day SJ, McDermott DF, Weber RW, Sosman JA, Haanen JB, et al. Improved survival with ipilimumab in patients with metastatic melanoma. N Engl J Med. 2010;363:711–23.

2. Topalian SL, Hodi FS, Brahmer JR, Gettinger SN, Smith DC, McDermott DF, et al. Safety, activity, and immune correlates of anti-PD-1 antibody in cancer. N Engl J Med. 2012;366:2443–54.

3. Brahmer JR, Tykodi SS, Chow LQM, Hwu W-J, Topalian SL, Hwu P, et al. Safety and Activity of Anti–PD-L1 Antibody in Patients with Advanced Cancer. N Engl J Med. 2012;366:2455–65.

4. Sharma P, Allison JP. The future of immune checkpoint therapy. Science. 2015;348:56–61.

5. Schumacher TN, Scheper W, Kvistborg P. Cancer Neoantigens. Annu Rev Immunol. 2019;37:173–200.

6. McLane LM, Abdel-Hakeem MS, Wherry EJ. CD8 T Cell Exhaustion During Chronic Viral Infection and Cancer. Annu Rev Immunol. 2019;37:457–95.

7. Zitvogel L, Ma Y, Raoult D, Kroemer G, Gajewski TF. The microbiome in cancer immunotherapy: Diagnostic tools and therapeutic strategies. Science. 2018;359:1366–70.

8. Flint TR, Janowitz T, Connell CM, Roberts EW, Denton AE, Coll AP, et al. Tumor-Induced IL-6 Reprograms Host Metabolism to Suppress Anti-tumor Immunity. Cell Metab. 2016;24:672–84.

9. Sahai E, Astsaturov I, Cukierman E, DeNardo DG, Egeblad M, Evans RM, et al. A framework for advancing our understanding of cancer-associated fibroblasts. Nat Rev Cancer. 2020;3:174–86.

10. Germain C, Gnjatic S, Tamzalit F, Knockaert S, Remark R, Goc J, et al. Presence of B cells in tertiary lymphoid structures is associated with a protective immunity in patients with lung cancer. Am J Respir Crit Care Med. 2014;189:832–44.

11. Petitprez F, de Reyniès A, Keung EZ, Chen TWW, Sun CM, Calderaro J, et al. B cells are associated with survival and immunotherapy response in sarcoma. Nature. 2020;577:556–60.

12. Helmink BA, Reddy SM, Gao J, Zhang S, Basar R, Thakur R, et al. B cells and tertiary lymphoid structures promote immunotherapy response. Nature. 2020;577:549–55.

13. Cabrita R, Lauss M, Sanna A, Donia M, Skaarup Larsen M, Mitra S, et al. Tertiary lymphoid structures improve immunotherapy and survival in melanoma. Nature. 2020;577:561–5.

14. Denton AE, Linterman MA. Stromal networking: cellular connections in the germinal centre. Curr Opin Immunol. 2017;45:103–111.

15. Denton AE, Carr EJ, Magiera LP, Watts AJB, Fearon DT. Embryonic FAP+ lymphoid tissue organizer cells generate the reticular network of adult lymph nodes. J Exp Med. 2019;216:2242–52.

16. Olumi AF, Grossfeld GD, Hayward SW, Carroll PR, Tlsty TD, Cunha GR. Carcinoma-associated fibroblasts direct tumor progression of initiated human prostatic epithelium. Cancer Res. 1999;59:5002–11.

17. Garin-Chesa P, Old LJ, Rettig WJ. Cell surface glycoprotein of reactive stromal fibroblasts as a potential antibody target in human epithelial cancers. Proc Natl Acad Sci U S A. 1990;87:7235–9.

18. Kraman M, Bambrough PJ, Arnold JN, Roberts EW, Magiera L, Jones JO, et al. Suppression of antitumor immunity by stromal cells expressing fibroblast activation protein-alpha. Science. 2010;330:827–30.

19. Orimo A, Gupta PB, Sgroi DC, Arenzana-Seisdedos F, Delaunay T, Naeem R, et al. Stromal fibroblasts present in invasive human breast carcinomas promote tumor growth and angiogenesis through elevated SDF-1/CXCL12 secretion. Cell. 2005;121:335–48.

20. Feig C, Jones JO, Kraman M, Wells RJB, Deonarine A, Chan DS, et al. Targeting CXCL12 from FAP-expressing carcinoma-associated fibroblasts synergizes with anti – PD-L1 immunotherapy in pancreatic cancer. Proc Natl Acad Sci U S A. 2013;110:20212–7.

21. Zhang WB, Navenot JM, Haribabu B, Tamamuraz H, Hiramatu K, Omagari A, et al. A point mutation that confers constitutive activity to CXCR4 reveals that T140 is an inverse agonist and that AMD3100 and ALX40-4C are weak partial agonists. J Biol Chem. 2002;277:24515–21.

22. Schall TJ, Proudfoot AEI. Overcoming hurdles in developing successful drugs targeting chemokine receptors. Nat Rev Immunol. 2011;11:355–63.

23. Hendrix CW, Collier AC, Lederman MM, Schols D, Pollard RB, Brown S, et al. Safety, pharmacokinetics, and antiviral activity of AMD3100, a selective CXCR4 receptor inhibitor, in HIV-1 infection. J Acquir Immune Defic Syndr. 2004;37:1253–62.

24. Peled A, Petit I, Kollet O, Magid M, Ponomaryov T, Byk T, et al. Dependence of human stem cell engraftment and repopulation of NOD/SCID mice on CXCR4. Science. 1999;283:845–8.

25. Gentles AJ, Newman AM, Liu CL, Bratman S V., Feng W, Kim D, et al. The prognostic landscape of genes and infiltrating immune cells across human cancers. Nat Med. 2015;21:938–45.

26. Siegert S, Luther SA. Positive and negative regulation of T cell responses by fibroblastic reticular cells within paracortical regions of lymph nodes. Front Immunol. 2012;3:285.

27. Denton AE, Roberts EW, Linterman MA, Fearon DT. Fibroblastic reticular cells of the lymph node are required for retention of resting but not activated CD8+ T cells. Proc Natl Acad Sci U S A. 2014;111:12139–12144.

28. Reeve J, Sellarés J, Mengel M, Sis B, Skene A, Hidalgo L, et al. Molecular diagnosis of T cell-mediated rejection in human kidney transplant biopsies. Am J Transplant. 2013;13:645–55.

29. Halloran PF, Pereira AB, Chang J, Matas A, Picton M, De Freitas D, et al. Potential impact of microarray diagnosis of t cell-mediated rejection in kidney transplants: The INTERCOM study. Am J Transplant. 2013;13:2352–63.

30. Kautto EA, Bonneville R, Miya J, Yu L, Krook MA, Reeser JW, et al. Performance evaluation for rapid detection of pan-cancer microsatellite instability with MANTIS. Oncotarget. 2017;8:7452–63.

31. Goldman M, Craft B, Kamath A, Brookes A, Zhu J, Haussler D. The UCSC Xena platform for cancer genomics data visualization and interpretation. bioRxiv. 2018;326470.

32. Riaz N, Havel JJ, Makarov V, Desrichard A, Urba WJ, Sims JS, et al. Tumor and Microenvironment Evolution during Immunotherapy with Nivolumab. Cell. 2017;171:934–949.e16.

33. Gide TN, Quek C, Menzies AM, Tasker AT, Shang P, Holst J, et al. Distinct Immune Cell Populations Define Response to Anti-PD-1 Monotherapy and Anti-PD-1/Anti-CTLA-4 Combined Therapy. Cancer Cell. 2019;35:238–55.

34. Griss J, Bauer W, Wagner C, Simon M, Chen M, Grabmeier-Pfistershammer K, et al. B cells sustain inflammation and predict response to immune checkpoint blockade in human melanoma. Nat Commun. 2019;10:4186.

35. Wan JCM, Massie C, Garcia-Corbacho J, Mouliere F, Brenton JD, Caldas C, et al. Liquid biopsies come of age: Towards implementation of circulating tumour DNA. Nat Rev Cancer. 2017;17:223–38.

36. Cabel L, Proudhon C, Romano E, Girard N, Lantz O, Stern MH, et al. Clinical potential of circulating tumour DNA in patients receiving anticancer immunotherapy. Nat Rev Clin Oncol. 2018;15:639–50.

37. Anagnostou V, Forde PM, White JR, Niknafs N, Hruban C, Naidoo J, et al. Dynamics of tumor and immune responses during immune checkpoint blockade in non–small cell lung cancer. Cancer Res. 2019;79:1214–25.

38. Parkinson CA, Gale D, Piskorz AM, Biggs H, Hodgkin C, Addley H, et al. Exploratory Analysis of TP53 Mutations in Circulating Tumour DNA as Biomarkers of Treatment Response for Patients with Relapsed High-Grade Serous Ovarian Carcinoma: A Retrospective Study. PLoS Med. 2016;13:e1002198.

39. Sanmamed MF, Carranza-Rua O, Alfaro C, Oñate C, Martín-Algarra S, Perez G, et al. Serum interleukin-8 reflects tumor burden and treatment response across malignancies of multiple tissue origins. Clin Cancer Res. 2014;20:5697–707.

40. Sanmamed MF, Perez-Gracia JL, Schalper KA, Fusco JP, Gonzalez A, Rodriguez-Ruiz ME, et al. Changes in serum interleukin-8 (IL-8) levels reflect and predict response to anti-PD-1 treatment in melanoma and non-small-cell lung cancer patients. Ann Oncol. 2017;28:1988–95.

41. Yuen KC, Liu LF, Gupta V, Madireddi S, Keerthivasan S, Li C, et al. High systemic and tumor-associated IL-8 correlates with reduced clinical benefit of PD-L1 blockade. Nat Med. 2020;26:693–698.

42. Hodi FS, Hwu W-J, Kefford R, Weber JS, Daud A, Hamid O, et al. Evaluation of Immune-Related Response Criteria and RECIST v1.1 in Patients With Advanced Melanoma Treated With Pembrolizumab. J Clin Oncol. 2016;34:1510–7.

43. Wolchok JD, Hoos A, O’Day S, Weber JS, Hamid O, Lebbé C, et al. Guidelines for the evaluation of immune therapy activity in solid tumors: immune-related response criteria. Clin Cancer Res. 2009;15:7412–20.

44. Bockorny B, Semenisty V, Macarulla T, Borazanci E, Wolpin BM, Stemmer SM, et al. BL-8040, a CXCR4 antagonist, in combination with pembrolizumab and chemotherapy for pancreatic cancer: the COMBAT trial. Nat Med. 2020;10.1038/s41591-020-0880-x.

45. Nayar S, Campos J, Smith CG, Iannizzotto V, Gardner DH, Mourcin F, et al. Immunofibroblasts are pivotal drivers of tertiary lymphoid structure formation and local pathology. Proc Natl Acad Sci U S A. 2019;116:13490–7.

46. Love MI, Huber W, Anders S. Moderated estimation of fold change and dispersion for RNA-seq data with DESeq2. Genome Biol. 2014;15:550.

47. Mouliere F, Chandrananda D, Piskorz AM, Moore EK, Morris J, Ahlborn LB, et al. Enhanced detection of circulating tumor DNA by fragment size analysis. Sci Transl Med. 2018;10:eaat4921.

